# Place, cause and expectedness of death and relationship to the deceased are associated with poorer experiences of end-of-life care and challenges in early bereavement: Risk factors from an online survey of people bereaved during the COVID-19 pandemic

**DOI:** 10.1101/2021.09.09.21263341

**Authors:** LE Selman, DJJ Farnell, M Longo, S Goss, K Seddon, A Torrens-Burton, CR Mayland, D Wakefield, B Johnston, A Byrne, E Harrop

## Abstract

**Objectives:** To identify clinical and demographic risk factors for sub-optimal end-of-life care and pandemic-related challenges prior to death and in early bereavement.

**Design:** Online open national survey of adults bereaved in the UK from 16 March 2020-5 January 2021, recruited via media, social media, national associations and organisations.

**Setting:** General population, UK.

**Participants:** 711 participants, mean age 49.5 (SD 12.9, range 18-90). 395 (55.6%) had experienced the death of a parent, 152 (21.4%) a partner. 628 (88.6%) were female and 33 (4.7%) from a minority ethnic background. The mean age of the person who died was 72.2 (SD 16.1, range miscarriage to 102 years). 311 (43.8%) deaths were from confirmed/suspected COVID-19, and 410 (57.8%) deaths occurred in hospital.

**Main outcome measures:** End-of-life care experiences (six items, e.g. involvement in care decisions) and pandemic-related challenges before and after death (six items, e.g. unable to visit prior to death).

**Results:** Deaths in hospital/care home increased the likelihood of: unable to visit prior to death, unable to say goodbye as wanted, limited contact in last days of life (all *P*<0.001). Deaths in hospice/at home increased the likelihood of: involved in care decisions (*P*<0.001), well supported by healthcare professionals (HCPs) after the death (*P*=0.003). Hospice deaths increased the likelihood of being given bereavement support information, which was least likely for care home deaths (*P*<0.001). Hospital deaths decreased the likelihood of knowing the contact details for the responsible care professional (*P*=0.001). Bereavement due to COVID-19 decreased the likelihood of: involvement in care decisions (*P*<0.001), feeling well supported by HCPs after the death (*P*<0.001), and increased the likelihood of: being unable to say goodbye (OR=0.348; 95% CI: 0.2 to 0.605), social isolation and loneliness (OR=0.439; 95% CI: 0.261 to 0.739), limited contact with relatives/friends (OR=0.465; 95% CI: 0.254 to 0.852). Expected deaths were associated with higher likelihood of feeling involved, informed, and well supported by HCPs (all *P*<0.001). The deceased being a partner or child increased the likelihood of knowing the contact details for the responsible care professional (*P*=0.001), being able to visit (*P*<0.001) and given bereavement support information (*P*<0.001). Being a bereaved partner strongly increased odds of social isolation and loneliness, e.g. OR = 0.092 (95% CI: 0.028 to 0.297) partner versus distant family member.

**Conclusions:** Four clear risk factors were found for poorer end-of-life care and pandemic-related challenges in bereavement: place, cause and expectedness of death, and relationship to the deceased.

What is already known on this topic?

- Since the start of the pandemic, over 20 million family members and friends have been bereaved due to COVID-19, with millions more bereaved due to other causes.
- Bereavement of any cause during the COVID-19 pandemic is associated with specific challenges, including limited access to people before their death, pressure on health and social care providers, quarantining due to infection or exposure, lockdowns and social distancing.
- There remains little evidence to inform optimal clinical practice, bereavement support and the policy response to COVID-19 as a mass bereavement event.

What this study adds

- Our study highlights four risk factors for poorer end-of-life care and increased risk of pandemic-related challenges in early bereavement: place, cause and expectedness of death and relationship to the deceased.
- COVID-19 deaths, hospital and care home deaths and unexpected deaths were generally associated with poorer outcomes, while being a partner of the person who died (regardless of cause) and bereavement due to COVID-19 increased the odds of experiencing social isolation and loneliness in bereavement.
- These factors should be taken into account in clinical practice, policy and bereavement support.

## Introduction

The COVID-19 pandemic has resulted in widespread, mass bereavement on an unprecedented global scale, with over 4 million deaths from COVID-19 recorded so far^1^. With each death associated with approximately nine close bereavements^2^, an estimated 20.8 million family members and friends have been bereaved due to COVID-19 since the start of the pandemic. Deaths during the pandemic are associated with risk factors for poor bereavement outcomes identified pre-pandemic, including traumatic end-of-life and death experiences, being unable to say goodbye, loss of community networks and social support, and social and economic disruption^3-9^. Many of these challenges are relevant to non-COVID-19 deaths as well as bereavements due to COVID-19.

End-of-life care and infection control restrictions such as social distancing policies have varied since the start of the pandemic, with inconsistencies across geographical areas and care settings. Recent research has described some of the challenges of providing end-of-life-care during the pandemic^10^ and qualitatively investigated bereavement experiences^11^ and support needs^12^. However, there remains little evidence to inform optimal clinical practice, bereavement support and the policy response to COVID-19 as a mass bereavement event. This study aimed to identify clinical and demographic risk factors associated with experiencing sub-optimal end-of-life care or pandemic-related challenges prior to death and in early bereavement, using baseline data from a mixed-methods longitudinal study of bereavement during the pandemic^12, 13^.

## Methods

### Design

An open web survey (Supplementary file 1) was designed by the research team, which includes a public representative (KS), with input from the study advisory group. It was piloted and refined with members of the public (see *Patient and Public Involvement*) and tested by the advisory group and departmental colleagues. Survey items and structure were informed by study aims and previous research^14-17^. Non-randomised open and closed questions covered end-of-life and grief experiences, and perceived needs for, access to and experiences of formal and informal bereavement support.

### Primary outcomes

#### Experiences of end-of-life care

Six items assessed end-of-life care experiences: involvement in decisions about the deceased’s care, knowing the contact details for the professional responsible for their care, receiving information about the approaching death, level of support by healthcare professionals immediately after the death, contact by the hospital or care provider after the death, and being provided with information about bereavement support services by the hospital or care provider. These questions were selected and adapted from the Consumer Quality Index for Palliative Care^14^ and used as quality markers of good end-of-life care.

#### Pandemic-related challenges

Six items assessed pandemic-related challenges prior to death and after the death, based on emerging evidence of the challenges of bereavement during the pandemic. Exploratory factor analysis found two subscales, related to problems due to “contact with loved one prior to the death” (namely: Were you unable to visit prior to death? Did you experience limited contact in last days of life? Were you unable to say goodbye?) and “social isolation” (namely: Did you experience restricted funeral arrangements? Did you experience any social isolation and loneliness? Did you experience limited contact with others?). All items were answered yes/no. For each participant, the numbers of problems experienced in each subscale and the total number of problems experienced over all six items were calculated by summing item scores (0=no and 1=yes; possible range: 0-3 for each subscale, 0-6 for total). Cronbach’s α was 0.57, 0.57, and 0.64 for the “contact prior to death” subscale (3 items), “social isolation” subscale (3 items) and total (all 6 items) respectively.

### Associated factors

We assessed whether demographic and clinical factors independently predicted end-of-life care experiences and pandemic-related challenges. Factors included in the analysis are recognised risk factors for poor bereavement outcomes (age, gender, relationship to deceased, expectedness of the death)^18, 19^ or have been identified as indirectly associated with experiences of end-of-life care (qualifications, deprivation level and region; place of death; cause of death)^20, 21^. We used postcode data to identify geographical region of residence and (for England) socio-economic deprivation.

### Study procedure

The survey was administered via JISC (https://www.onlinesurveys.ac.uk/) and was open from 28th August 2020 to 5th January 2021. It was disseminated to a convenience sample on social and mainstream media and via voluntary sector associations and bereavement support organisations, including community and national organisations representing ethnic minority communities. These organisations helped disseminate the voluntary survey by sharing on social media, web-pages, newsletters, on-line forums and via direct invitations to potential participants (see Supplementary file 2 for an example advertisement). For ease of access, the survey was posted onto a bespoke study-specific website with a memorable URL (covidbereavement.com). Two participants chose to complete the survey in paper format. Summaries of survey results (including interim results released November 2020) were posted on the study website and provided to interested participants.

Inclusion criteria: aged 18+; family or close friend bereaved since social-distancing requirements were introduced in the UK (16/03/2020); death occurred in the UK; ability to consent. The initial section of the survey requested informed consent and details of data protection (see Supplementary file 1).

Reporting follows the Checklist for Reporting Results of Internet E-Surveys^22^.

### Patient and public involvement

Co-author KS is a public representative with experience of bereavement. She was a co-applicant on the funding application and is a member of the research team, contributing at all stages of the research. The survey and recruitment strategy were developed through consultation with patients and public representatives at Cardiff University and the University of Bristol. The survey was piloted by 16 public representatives with experience of bereavement, with wording and content refined based on their feedback. A plain English summary of this paper will be produced with public representatives and emailed to study participants, made available on the study website (covidbereavement.com) and disseminated via social media.

### Data analysis

Frequency tables were used to explore the data initially. Cohen’s *h* effect size effect was used to measure the differences between two proportions. For any comparisons, the maximum value of *h* is presented to provide an estimate of (maximum) effect size (*h* = 0.2: small effect, *h* = 0.5: medium effect, *h* ≥ 0.8: large effect). Chi-squared tests and Fisher’s exact test were used for categorical outcome data. Mixed-effects logistic regression implemented via mixed-effects generalised linear model (GLM) was used to examine pandemic-specific challenges as a function of factors identified as having a strong and / or significant effect on outcomes, as derived from Cohen’s *h* effect sizes and univariate calculations. A Directed Acyclic Graph (DAG) was also used to visualise the relationships between variables. Note that region of the UK was therefore included as a random effect, whereas all other variables were coded as fixed effects. Results of the mixed-effects GLM analysis are presented as odds ratios in supplementary tables. To provide a comparison to the results from the mixed-effects GLM, odds ratios were also found via simple logistic regression. All calculations were carried out using SPSS V26.

## Results

### Sample characteristics

711 bereaved participants completed the survey (Table 1). 12 surveys were completed in duplicate and were screened out using contact and demographic information, with the first completed survey retained for these participants. Two incomplete surveys were excluded where only the consent question had been answered. Missing data was minimal for all variables, with an average amount of missing data per item of 0.7% (range 0-3.7%). Imputation of missing data was not necessary.

**Table 1.**
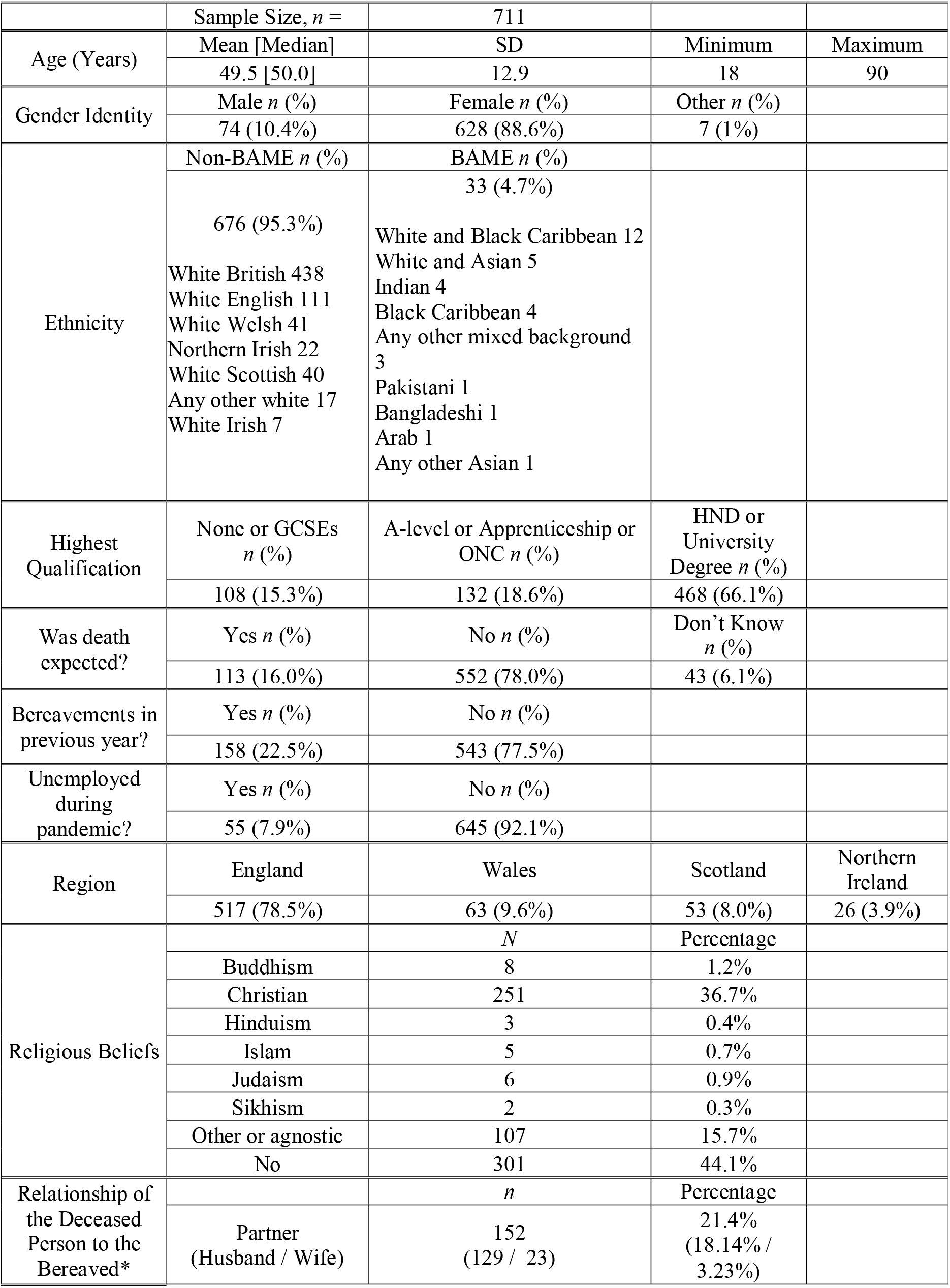
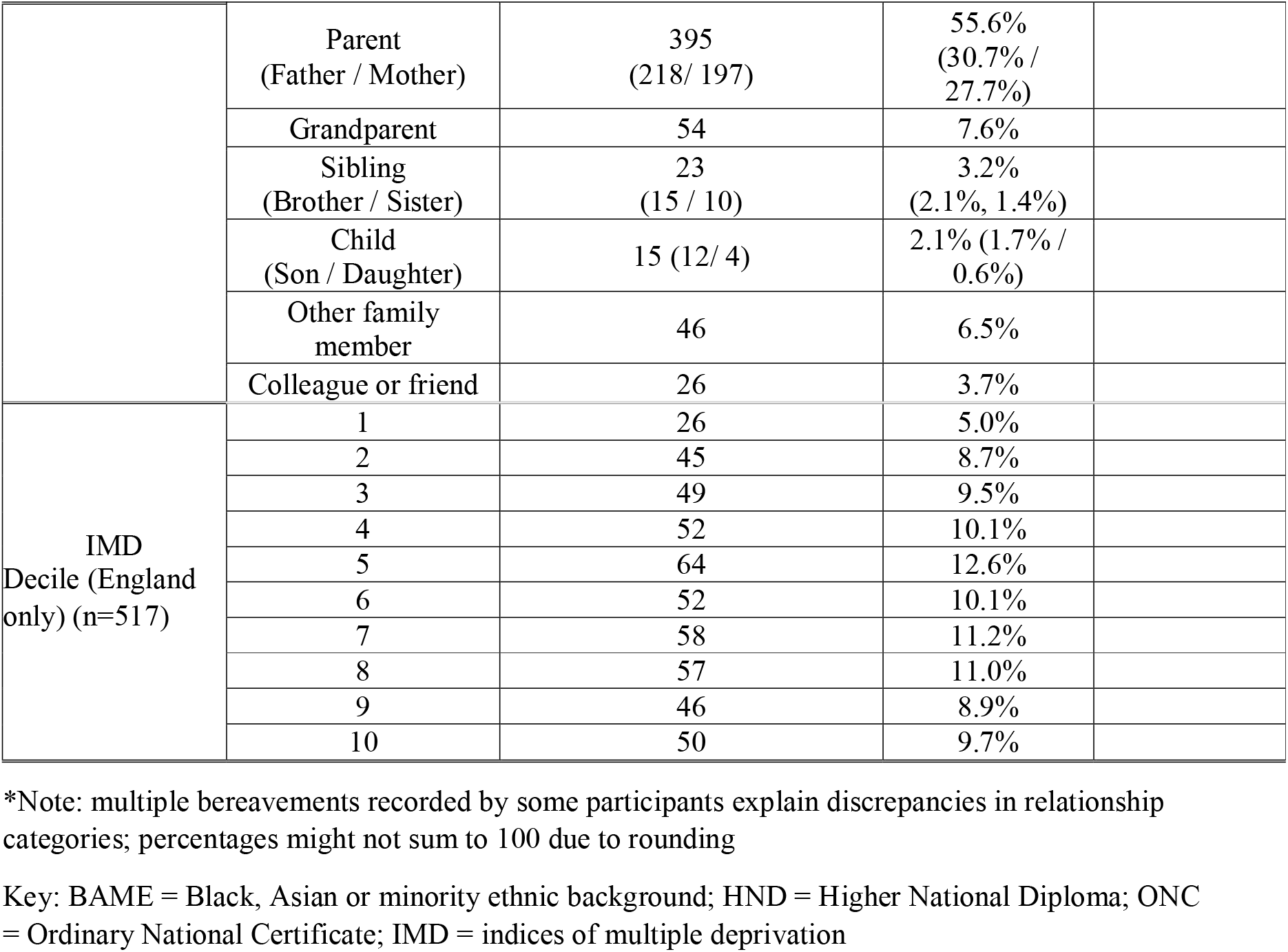
Characteristics of the bereaved.

|  |  |  |  |  |
| --- | --- | --- | --- | --- |
|  | Sample Size, *n* = | 711 |  |  |
| Age (Years) | Mean [Median] | SD | Minimum | Maximum |
|  | 49.5 [50.0] | 12.9 | 18 | 90 |
| Gender Identity | Male *n* (%) | Female *n* (%) | Other *n* (%) |  |
|  | 74 (10.4%) | 628 (88.6%) | 7 (1%) |  |
| Ethnicity | Non-BAME *n* (%) | BAME *n* (%) |  |  |
|  | 676 (95.3%)<br>White British 438<br>White English 111<br>White Welsh 41<br>Northern Irish 22<br>White Scottish 40<br>Any other white 17<br>White Irish 7 | 33 (4.7%)<br>White and Black Caribbean 12<br>White and Asian 5<br>Indian 4<br>Black Caribbean 4<br>Any other mixed background 3<br>Pakistani 1<br>Bangladeshi 1<br>Arab 1<br>Any other Asian 1 |  |  |
| Highest Qualification | None or GCSEs *n* (%) | A-level or Apprenticeship or ONC *n* (%) | HND or University Degree *n* (%) |  |
|  | 108 (15.3%) | 132 (18.6%) | 468 (66.1%) |  |
| Was death expected? | Yes *n* (%) | No *n* (%) | Don't Know *n* (%) |  |
|  | 113 (16.0%) | 552 (78.0%) | 43 (6.1%) |  |
| Bereavements in previous year? | Yes *n* (%) | No *n* (%) |  |  |
|  | 158 (22.5%) | 543 (77.5%) |  |  |
| Unemployed during pandemic? | Yes *n* (%) | No *n* (%) |  |  |
|  | 55 (7.9%) | 645 (92.1%) |  |  |
| Region | England | Wales | Scotland | Northern Ireland |
|  | 517 (78.5%) | 63 (9.6%) | 53 (8.0%) | 26 (3.9%) |
| Religious Beliefs |  | *N* | Percentage |  |
|  | Buddhism | 8 | 1.2% |  |
|  | Christian | 251 | 36.7% |  |
|  | Hinduism | 3 | 0.4% |  |
|  | Islam | 5 | 0.7% |  |
|  | Judaism | 6 | 0.9% |  |
|  | Sikhism | 2 | 0.3% |  |
|  | Other or agnostic | 107 | 15.7% |  |
|  | No | 301 | 44.1% |  |
| Relationship of the Deceased Person to the Bereaved* |  | *n* | Percentage |  |
|  | Partner (Husband / Wife) | 152 (129 / 23) | 21.4% (18.14% / 3.23%) |  |

|  |  |  |  |
| --- | --- | --- | --- |
|  | Parent<br>(Father / Mother) | 395<br>(218/ 197) | 55.6%<br>(30.7% /<br>27.7%) |
|  | Grandparent | 54 | 7.6% |
|  | Sibling<br>(Brother / Sister) | 23<br>(15 / 10) | 3.2%<br>(2.1%, 1.4%) |
|  | Child<br>(Son / Daughter) | 15 (12/ 4) | 2.1% (1.7% /<br>0.6%) |
|  | Other family<br>member | 46 | 6.5% |
|  | Colleague or friend | 26 | 3.7% |
| IMD<br>Decile (England<br>only) (n=517) | 1 | 26 | 5.0% |
|  | 2 | 45 | 8.7% |
|  | 3 | 49 | 9.5% |
|  | 4 | 52 | 10.1% |
|  | 5 | 64 | 12.6% |
|  | 6 | 52 | 10.1% |
|  | 7 | 58 | 11.2% |
|  | 8 | 57 | 11.0% |
|  | 9 | 46 | 8.9% |
|  | 10 | 50 | 9.7% |

\*Note: multiple bereavements recorded by some participants explain discrepancies in relationship categories; percentages might not sum to 100 due to rounding
Key: BAME = Black, Asian or minority ethnic background; HND = Higher National Diploma; ONC = Ordinary National Certificate; IMD = indices of multiple deprivation

Participants represented diverse geographical areas, deprivation indexes and levels of education. 628 (88.6%) of participants were female; the mean age of the bereaved person was 49.5 years old (SD = 12.9; range 18-90). 395 (55.6%) of participants had experienced the death of a parent, followed by partner/spouse (*n =* 152, 21.4%). 72 people (10.1%) had experienced more than one bereavement since 16^th^ March 2020. 33 people (4.7%) self-identified as from a minority ethnic background.

The mean age of the deceased person was 72.2 years old (SD=16.1; range: miscarriage at 4 months to 102 years’ old) (Table 2). 311 (43.8%) died of confirmed/suspected COVID-19, 156 (21.9%) from cancer, and 119 (16.7%) from another life-limiting condition. Most died in hospital (*n =* 410; 57.8%). Questionnaires were completed a median of 152 days (5 months) after the death (range 1-279 days).

**Table 2:**
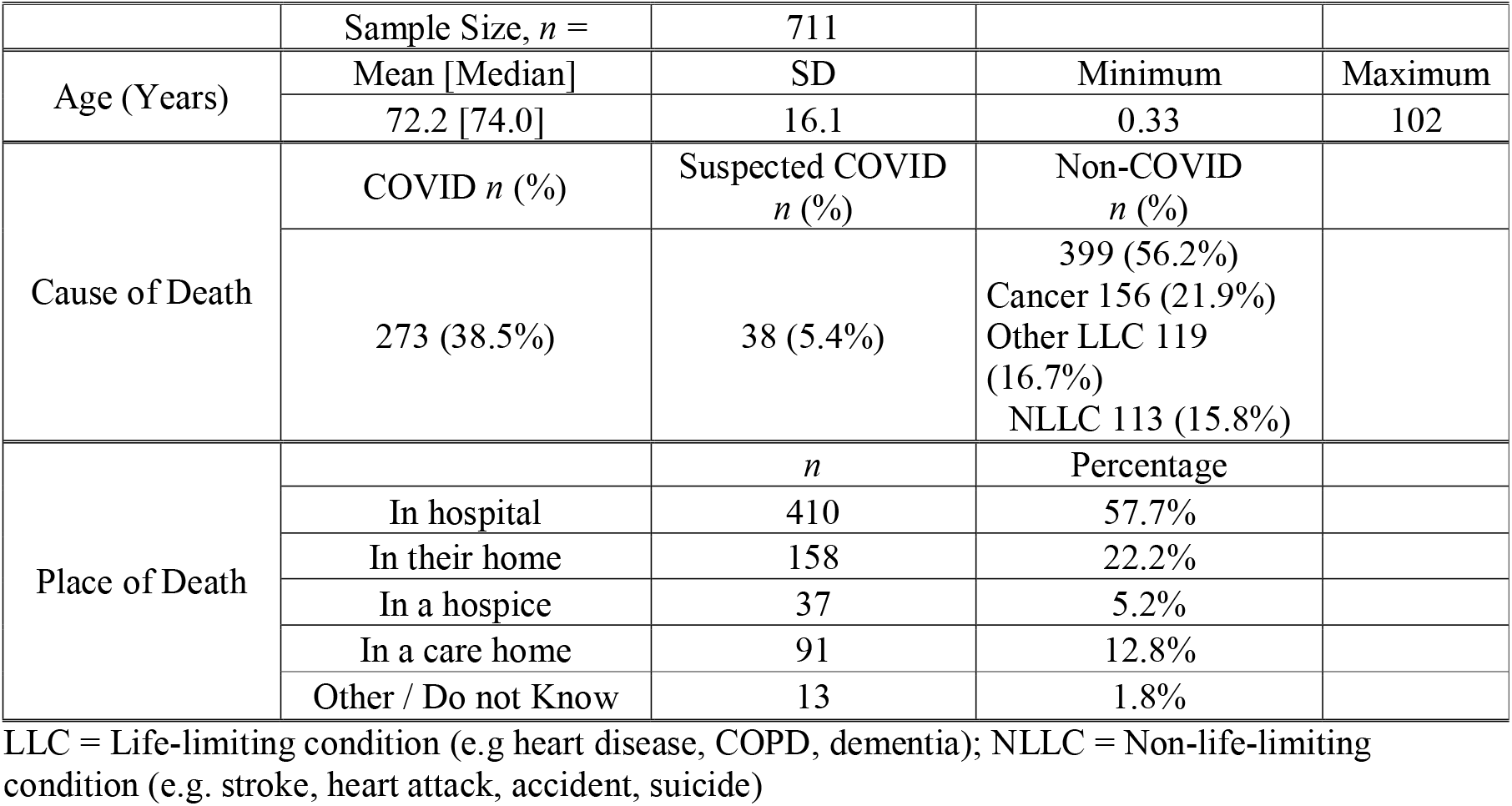
Characteristics of the deceased.

|  |  |  |  |  |
| --- | --- | --- | --- | --- |
|  | Sample Size, *n* = | 711 |  |  |
| Age (Years) | Mean [Median] | SD | Minimum | Maximum |
|  | 72.2 [74.0] | 16.1 | 0.33 | 102 |
| Cause of Death | COVID *n* (%) | Suspected COVID<br>*n* (%) | Non-COVID<br>*n* (%) |  |
|  | 273 (38.5%) | 38 (5.4%) | 399 (56.2%)<br>Cancer 156 (21.9%)<br>Other LLC 119<br>(16.7%)<br>NLLC 113 (15.8%) |  |
| Place of Death |  | *n* | Percentage |  |
|  | In hospital | 410 | 57.7% |  |
|  | In their home | 158 | 22.2% |  |
|  | In a hospice | 37 | 5.2% |  |
|  | In a care home | 91 | 12.8% |  |
|  | Other / Do not Know | 13 | 1.8% |  |

LLC = Life-limiting condition (e.g heart disease, COPD, dementia); NLLC = Non-life-limiting condition (e.g. stroke, heart attack, accident, suicide)

### Main outcomes

#### End-of-life care experiences

There was wide variation in overall reported experiences of end-of-life care (Table 3); for example, while 21.8% reported they were always involved in decision about the care of their loved one, 21.8% reported that they were never involved; 32.3% reported that they were fully informed about the approaching death while 17.7% said they were not at all informed. Half of the sample (49.8%) knew the contact details for the professional responsible for their loved one’s care. Over a quarter (28.2%) reported that they were very or fairly well supported by professionals immediately after the death, while 35.4% felt not at all supported. Overall, a third (34%) reported that a healthcare or other care professional had provided information about bereavement support services. Between 11.7% and 19.7% of respondents answered ‘not relevant’ to these questions e.g. because they were not next of kin, or no healthcare providers were involved in the death.

**Table 3:** Frequency of end-of-life care experiences.

|  |  | N | % |
| --- | --- | --- | --- |
| **Did the care professionals involve you in decisions about the care for your sick loved one?** | Never | 155 | 21.8 |
|  | Sometimes | 162 | 22.8 |
|  | Usually | 98 | 13.8 |
|  | Always | 155 | 21.8 |
|  | Not relevant to my situation (e.g. not next of kin, because none were involved) | 140 | 19.7 |
|  | Missing | 1 | 0.1 |
| **Did you know the contact details for the professional responsible for their care?** | Yes | 354 | 49.8 |
|  | No | 193 | 27.1 |
|  | Not sure | 52 | 7.3 |
|  | Not relevant to my situation | 109 | 15.3 |
|  | Missing | 3 | .4 |
| **Did you receive information about the approaching death?** | No, not at all | 126 | 17.7 |
|  | A bit of information | 270 | 38.0 |
|  | Yes, I was fully informed | 230 | 32.3 |
|  | Not relevant to my situation | 83 | 11.7 |
|  | Missing | 2 | .3 |
| **Did you feel well supported by the healthcare professionals immediately after the death of your loved one?** | Very well supported | 95 | 13.4 |
|  | Fairly well supported | 105 | 14.8 |
|  | A little bit supported | 139 | 19.5 |
|  | Not at all supported | 252 | 35.4 |
|  | Not relevant to my situation (e.g. because none were involved or not next of kin) | 120 | 16.9 |
| **Were you contacted again by the hospital or care provider following their death?** | Yes | 251 | 35.3 |
|  | No | 322 | 45.3 |
|  | Not relevant to my situation | 138 | 19.4 |
| **Did they provide information about bereavement support services?** | Yes (at the time of death) | 131 | 18.4 |
|  | Yes (during follow up call) | 89 | 12.5 |
|  | Yes (at the time of death and during follow up call) | 22 | 3.1 |
|  | No | 342 | 48.1 |
|  | Not relevant to my situation | 119 | 16.7 |
|  | Missing | 8 | 1.1 |

#### Pandemic-related challenges

Bereaved participants experienced a mean of 4.17 (median = 4) different types of pandemic-specific problems (out of a maximum of 6) (Table 4). People reported significantly higher levels of problems due to social isolation (mean = 2.41, median = 3) than problems related to contact before death (mean = 1.76, median = 2; Wilcoxon signed-rank test: *z* = 12.344, *P* < 0.001). The three most prevalent items were restricted funeral arrangements (93.4%), limited contact with other close relatives or friends (80.7%) and social isolation and loneliness (66.7%).

**Table 4:** Frequency of pandemic-related challenges by the bereaved before or after the death.

| **Subscale** | **Item** | **Percentage (95% CI)\*** |
| --- | --- | --- |
| Contact prior to death | Unable to visit them prior to their death | 54.3% (50.5% to 58.0%) |
|  | Limited contact with them in last days of their life | 57.8% (54.1% to 61.5%) |
|  | Unable to say goodbye as I would have liked | 63.9% (60.2% to 67.4%) |
| Social isolation | Restricted funeral arrangements | 93.4% (91.3% to 95.1%) |
|  | Social isolation and loneliness | 66.7% (63.1% to 70.1%) |
|  | Limited contact with other close relatives or friends | 80.7% (77.6% to 83.6%) |

\*Note that percentages are with respect to those participants who responded “yes” to these items.

### Place of death

Place of death had a moderate to strong influence on end-of-life care outcomes (Tables S1 to S7). If the death occurred in a hospice or at home the bereaved were more likely to be involved in decisions about the care for their loved one (*P* < 0.001) and feel well supported by the healthcare professionals immediately after the death of their loved one (*P* = 0.003) than if they had died in a hospital or care home (Table S1 and S4). If the person had died in hospital the bereaved was less likely to know the contact details for the professional responsible for their loved one’s care (*P* = 0.001) compared with other settings. Where the death had occurred in a hospice the bereaved person was most likely to have been provided with information about bereavement support services both at the time of death and during a follow-up call, while this was least likely for care home deaths (*P* < 0.001).

The effect size for place of death was high across all pandemic-related challenges. When a person had died in hospital or in a care home participants were most likely to report problems for all items in the “contact with loved one prior to death” subscale (*P* < 0.001) compared with other places of death (Table 5). Mixed-effects GLM analyses showed clearly that when death occurred at home, in a hospice or “other / don’t know” the bereaved participant had strongly decreased odds of being unable to visit their loved one before death, limited contact with them in last days of their life and being unable to say goodbye as they would have liked compared with death in hospital (Tables S8 to S10). Differences did not reach statistical significance for restricted funeral arrangements, social isolation and loneliness, and limited contact with other close relatives or friends (Table 5; Tables S11 to S13).

**Table 5:**
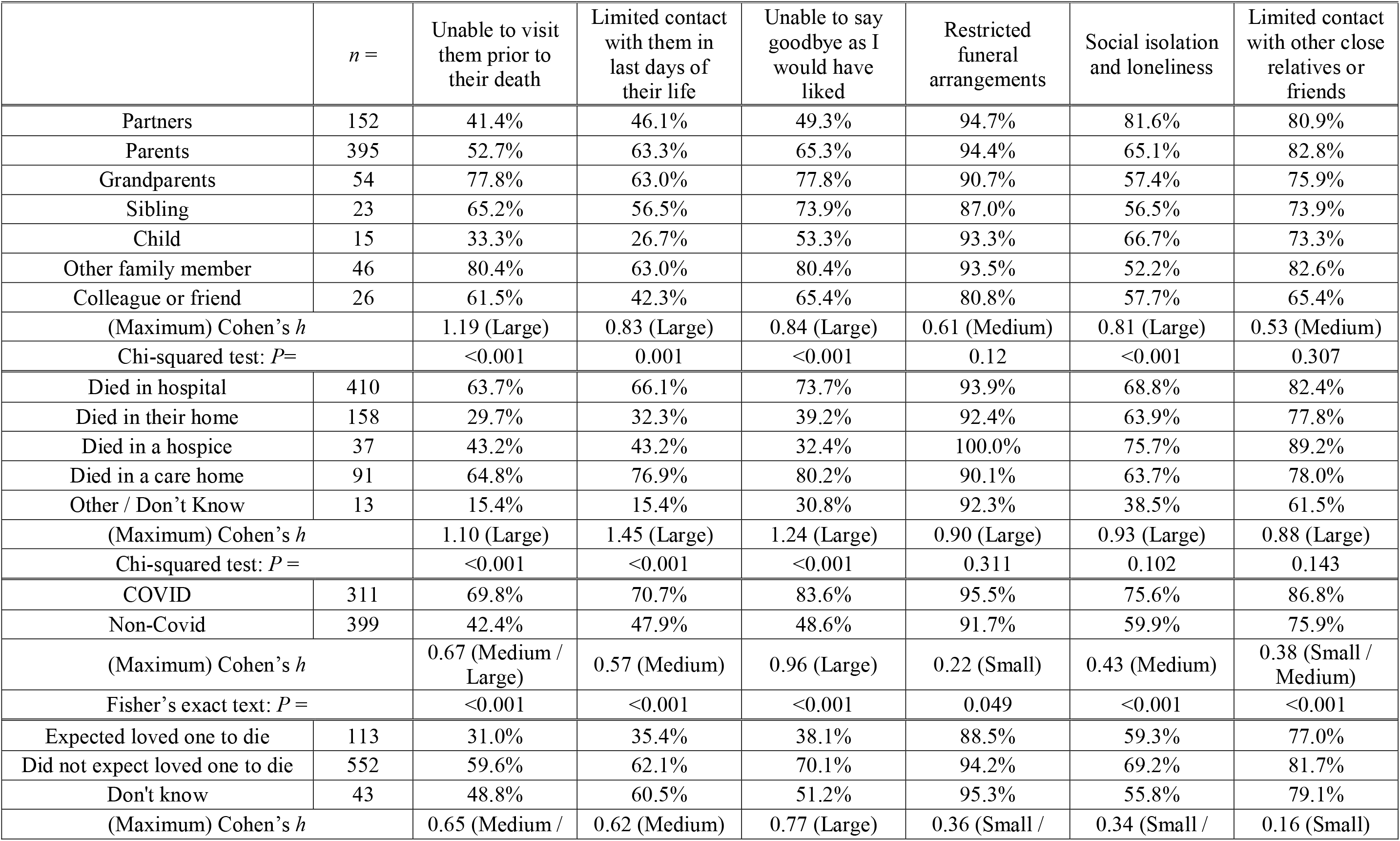
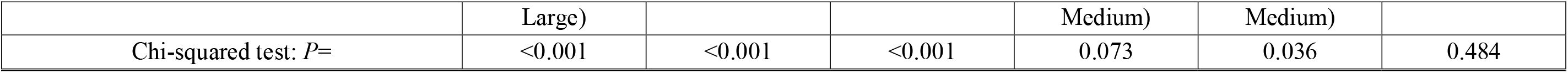
Pandemic-related challenges compared by: relationship to deceased, place of death, cause of death and unexpectedness of death. Note that percentages are with respect to those participants who responded “yes” to the pandemic-related challenge items. Effect sizes are estimated from the maximum Cohen’s *h* between any two groups for a given factor, where: *h* = 0.2: small effect, *h* = 0.5: medium effect, *h* ≥ 0.8: large effect.

### Cause of death

Cause of death (COVID versus non-COVID) had a moderate to weak effect on end-of-life care outcomes (Tables S1 to S7), with deaths due to COVID-19 associated with worse outcomes (Figure 1). In particular, participants bereaved due to COVID-19 were less likely to be involved in care decisions (*P* < 0.001) (Table S1) and less likely to be well supported by the healthcare professionals immediately after the death (*P* < 0.001) (Table S4).

**Figure 1:**
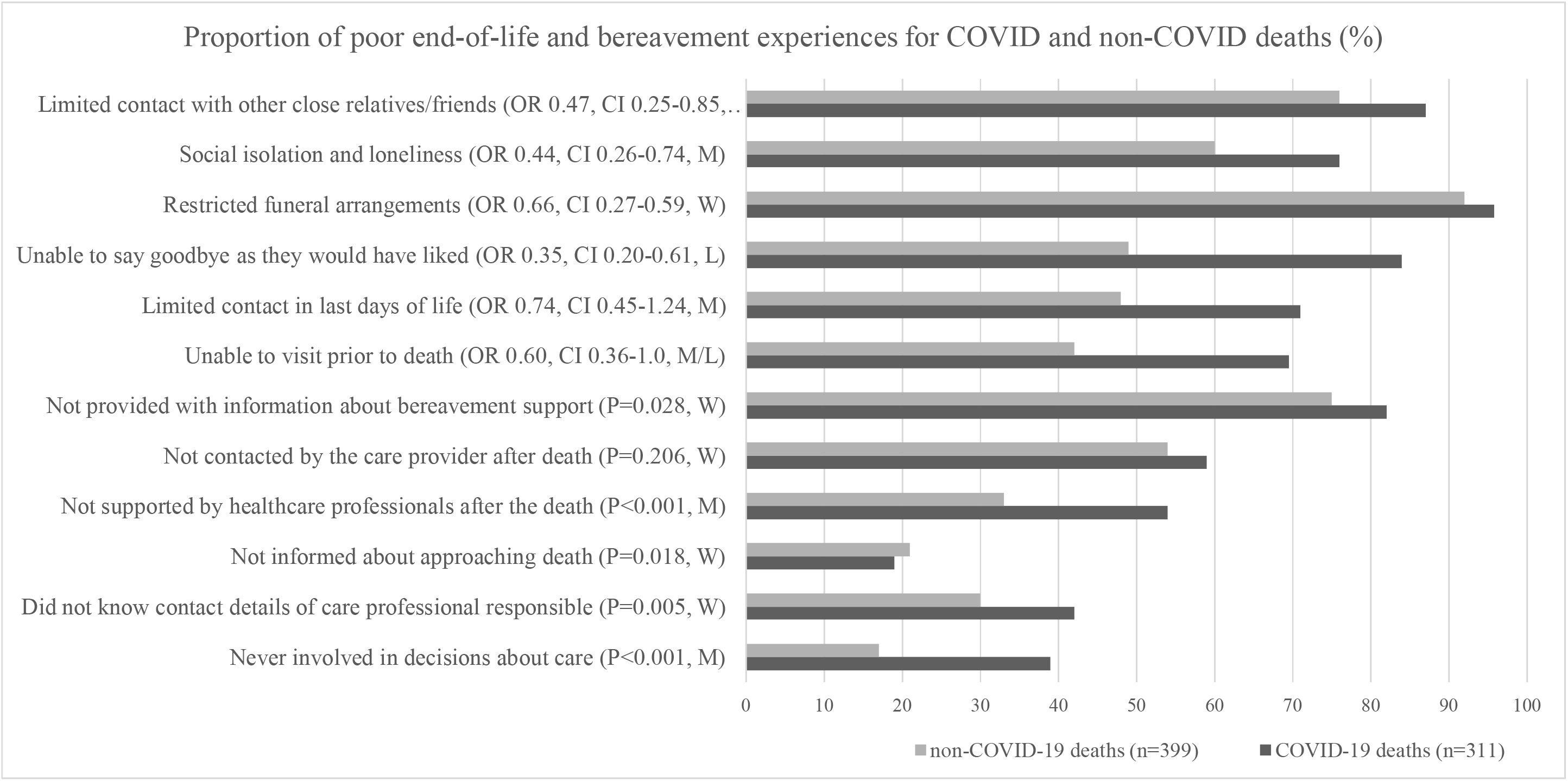
Proportion of poor end-of-life and bereavement experiences for COVID-19 and non-COVID deaths (%) Note: OR is for mixed-effects generalised linear model; 95% CIs; W=weak effect (Cohen’s *h* = 0.2); M=medium effect (Cohen’s *h* = 0.5); L=large effect (Cohen’s *h* = 0.8)

COVID-19 deaths were also associated with worse pandemic-related challenges, with weak to large effects sizes (Table 5, Figure 1). The total number of types of pandemic-related challenges was higher for COVID deaths compared to non-COVID (P < 0.001) (Table 5), and there was a consistent picture of increased odds of experiencing challenges (Tables S8 to S13; Figure 1). In particular, significant increases in odds were seen for: unable to say goodbye as I would have liked (Table S10, OR = 0.348; 95% CI: 0.2-0.605); social isolation and loneliness (Table S12, OR = 0.439; 95% CI: 0.261-0.739); and limited contact with other close relatives or friends (Table S13, OR = 0.465; 95% CI: 0.254-0.852).

### Expectedness of the death

Whether the death was expected or not had a moderate to strong effect on four end-of-life care outcomes (Tables S1 to S4), namely: an expected death led to the bereaved being more likely to be involved in decisions about the care for their loved one, know the contact details for the professional responsible for their care, receive information about their approaching death, and be well supported by the healthcare professionals immediately after the death (all *P* < 0.001).

The bereaved person expecting their loved one to die was also significantly associated with fewer experiences of some pandemic-related challenges (often *P* < 0.001) (Table 5). Results of logistic regression supported these results, with a consistent picture of decreased odds for expected deaths, however these findings were not confirmed by the GLM (Tables S8 to S13).

### Relationship to the deceased

Closer relationships (especially the deceased being a partner or child – also parents/siblings in some cases) compared with more distant relationships (distant family / colleague or friend) led to the bereaved person being more likely to know the contact details for the professional responsible for their loved one’s care (*P* = 0.001) and to be provided with information about bereavement support services at the time of death (*P* < 0.001). The relationship of the deceased to the bereaved had a medium to large effect on these variables (Tables S1 to S7.)

The relationship of the bereaved to the deceased person was also significantly associated with some of the pandemic-specific challenges. The inability to visit prior to death was highest if the deceased was a grandparent (77.8%) and lowest if the deceased was their child (33.3%) or partner (41.4%) compared to other groups (*P* < 0.001) (Table 5). Social isolation and loneliness was highest for bereaved partners (81.6%) compared with other groups (*P* < 0.001).

Results from simple logistic regression and mixed-effects GLM agree with these analyses. Relationships other than ‘partner’ had strongly increased odds of being unable to visit their loved one before death, e.g., participants whose grandparent had died were 9 times more likely not to have been able to visit them before death compared with bereaved partners (OR = 9.332, 95% CI: 2.033-42.841) (Table S8). Similar patterns occurred for limited contact with them in last days of their life and being unable to say goodbye (Tables S9 and S10). Parents whose child had died had a decreased risk of limited contact with them in last days of their life compared with partners (GLM: OR = 0.094; 95% CI: 0.009-0.982) (Table S10). Odds of restricted funeral arrangements were reduced for all groups compared with the reference group of partner, although this was significant only for colleague or friend versus partner (OR = 0.233; 95% CI: 0.070-0.781) (Table S11). The odds of social isolation and loneliness were strongly and significantly reduced for all groups compared to the reference group of partner, e.g., OR = 0.092 (95% CI: 0.028-0.297) for other family member (Table S12). Odds of limited contact with other close relatives or friends were not significantly different (Table S13).

### Demographic characteristics

#### Qualifications

Bereaved participants with higher levels of qualification were significantly more likely to be well supported by healthcare professionals immediately after the death (*P* = 0.028), and be contacted again by the hospital or care provider following the death (*P* = 0.011) (Tables S1 to S8).

#### Deprivation and region of the UK

Decile of deprivation (England) and region of the UK occasionally had a strong or moderate effect on end-of-life care outcomes (Tables S1 to S8). However, there were no significant differences (*P* > 0.05) and no obvious and consistent pattern across outcomes (region of the UK was therefore included in the mixed-effects GLM as a random-effect.)

#### Gender

The effects of gender identity were generally weak (Tables S1 to S7). Odds of social isolation and loneliness and separately limited contact with other close relatives or friends were higher for women compared with men, although this was only significant for limited contact with other close relatives or friends. The odds of restricted funeral arrangements were higher for women compared with men, although this was significant only for simple logistic regression (Table 13, GLM: OR = 2.496; 95% CI = 1.185 to 5.255).

#### Other demographic factors

Age of the deceased and the bereaved, religious belief, ethnicity, time since death, qualification level and same/different sex partnership had small effects on outcomes, none of which were significant (data not shown).

## Discussion

Place, cause and expectedness of death and relationship with the deceased were risk factors for sub-optimal end-of-life care and challenging experiences in early bereavement, reflecting pressures on health and social care settings and providers during the COVID-19 pandemic. Place of death had a moderate to strong effect on end-of-life care outcomes. For example, if the death occurred in a hospice or at home the bereaved were more likely to be involved in decisions about the care of their loved one and feel well supported by the healthcare professionals immediately after their death than if they had died in a hospital or care home. A bereaved person was least likely to have been provided with information about bereavement support services when the death had occurred in a care home. Hospice deaths and, as one would expect, home deaths were associated with fewer problems related to contact with the patient prior to death compared with hospital and care home deaths. Odds of social isolation and loneliness in early bereavement were highest for hospital deaths compared with other settings. Deaths due to COVID-19 were moderately to weakly associated with worse experiences of end-of-life care and perceived professional support after the death. People bereaved by COVID-19 were also particularly impacted by pandemic-related challenges, such as being unable to visit their loved one or say goodbye as they would have liked, limited contact with other relatives or friends, and social isolation and loneliness. This could reflect quarantining requirements after being close to an infected loved one, but also lockdown restrictions, fear and anxiety around social mixing and catching or spreading the virus, and the relationship challenges and sense of alienation associated with COVID-19 bereavement^12, 23, 24^.

Expectedness of death had a moderate effect on end-of-life care outcomes. An unexpected death led to the bereaved being less likely to be involved in care or, as one would expect, receive information prior to the death; however people experiencing a unexpected death were also less likely to feel supported by healthcare professionals or be contacted again. People who did not expect their loved one to die also had slightly increased odds of social isolation and loneliness in bereavement.

Partners and parents of the deceased were more likely to know the professional responsible for their loved one’s care, be provided with information about bereavement support, and be able to visit their loved one before the death compared with more distant family members or friends. However, social isolation and loneliness was highest among partners compared with other groups. There was evidence that bereaved people with higher levels of qualification received better support from healthcare professionals and more contact following the death, possibly because people with higher levels of education are more able to elicit support from healthcare professionals. This needs exploration in future research. Compared to men, women seemed more likely to report limited contact with other close relatives or friends; this also warrants further research.

This is the first study to identify risk factors for poor experiences among people bereaved during the COVID-19 pandemic. The sample was large, with good spread across geographical areas, education and deprivation, but was biased towards female and white respondents, despite targeted advertising to men and people from ethnic minority communities. By recruiting mostly online, we were less likely to reach the very old or other digitally marginalised groups, hence the high levels of social isolation we identified might under-estimate levels in the general bereaved population. Convenience sampling might also have resulted in more people with negative experiences completing the survey. Despite these limitations, group sizes were sufficient to enable comparisons (although not to the level of specific ethnic groups) and, while not providing population-level prevalence data, the sample does enable, for the first time, identification of risk factors to inform future practice and policy.

Bereaved people reported worse experiences in relation to hospital and care home deaths than deaths at home or hospice, as in pre-pandemic studies^25-28^. In the first ten weeks of the pandemic in the UK, deaths in care homes increased by 220%, and home and hospital deaths by 77% and 90%, respectively, while hospice deaths fell by 20%^29^. The increase in home deaths was sustained^30^ and hospices shifted their resources to the community^31^. Our findings suggest that despite the rise in home deaths during the pandemic, they were associated with better experiences of end-of-life care than deaths in other settings, indicating that primary and community care services were successful in supporting home deaths, particularly in light of the additional pressures on services^32^.

The finding that COVID-19 deaths were associated with poorer end-of-life and early bereavement experiences lends some support to the hypothesis that the pandemic will increase levels of prolonged grief disorder and other longer-term poor bereavement outcomes. Among the COVID-19 bereaved, such outcomes might be explained by the higher likelihood of poor end-of-life care experiences when a death is unexpected as well as the increased likelihood of pandemic-related challenges due to infection control restrictions. The current study’s longitudinal and qualitative data will throw further light on outcomes and experiences in this sample.

We found increased levels of social isolation and loneliness among people bereaved due to COVID-19, with bereaved partners at particular risk. In contrast, a survey in the Netherlands^33^ found that satisfaction with social support did not differ between people experiencing deaths caused by COVID-19 versus other types of deaths – however, given the small number of COVID-19 deaths (*n* = 49) these findings should be treated with caution. A US survey of people bereaved from COVID-19 (*n* = 307) found a close relationship with the deceased was associated with reduced functional impairment due to the loss^34^. In China (*n* = 422), the death of a close relation (partner, child or parent) due to COVID-19 was associated with more severe grief symptoms^35^. The higher levels of loneliness and social isolation observed amongst bereaved partners in this study may help explain these associations.

The evidence of sub-optimal end-of-life care demonstrates the difficulty of adequately supporting families during the pandemic, but also highlights areas for improvement. Communication with relatives must be prioritised and contact with loved ones at the end of life facilitated and optimised, even in the context of a pandemic. It is therefore crucial that end-of-life-care providers are prioritised when supplies of personal protective equipment (PPE) are overstretched, so that they are able to offer in-person visits, however there is evidence that this did not occur in 2020^36^. When patients are admitted to hospital with COVID-19, an unpredictable clinical course is likely and the risk of sudden death high. Given the risks associated with unexpected deaths, discussions with next of kin should happen early, with the risk of death explained clearly and compassionately. Partners bereaved due to an unexpected COVID-19 death in hospital may be at particular risk of poor outcomes and need additional follow-up and support, particularly when they live alone or have a previous history of mental disorders^37^. However, many challenges were experienced by people bereaved by non-COVID-19 deaths, and difficulties across the bereaved population should not be minimised. In particular, after sudden deaths of any cause bereaved relatives require attention and follow-up, given perceptions of poor support after death. Clear and consistent national guidance on hospital, hospice and care home visiting is essential to ensure equity and support staff. We found only a third of bereaved people had been given information about bereavement support services. Signposting at the time of death and in follow-up must be improved across settings to ensure bereaved people know how and where to seek support and help alleviate barriers to people accessing support^38^.

Further research is needed to examine the impact of end-of-life care experiences and pandemic-related social challenges on bereavement outcomes, and to determine the prevalence of poor bereavement outcomes including prolonged grief disorder among people bereaved during the pandemic, with comparisons to non-pandemic times. This is especially crucial given some discrepancies in emerging evidence^39-41^. The experiences of bereaved men and people from black and minority ethnic communities during the pandemic require further research.

## Conclusions

This online survey of more than 700 people bereaved during the pandemic found place, cause and expectedness of deaths and relationship to the deceased were risk factors for sub-optimal end-of-life care and challenging experiences in early bereavement during the pandemic. People bereaved by COVID-19 and partners bereaved by all causes of death were particularly at risk of social isolation and loneliness. To learn from COVID-19 as a mass bereavement event, these findings should inform optimal clinical practice, bereavement support and the policy response.

## Supporting information

CHERRIES reporting checklist

## Data Availability

Full study data sets will be made available following study closure in February 2022. Data sharing requests will be considered prior to this and should be directed to Dr Emily Harrop.

## Ethics approval

The study protocol and supporting documentation was approved by Cardiff University School of Medicine Research Ethics Committee (SMREC 20/59). The study was conducted in accordance with the Declaration of Helsinki and all respondents provided informed consent.

## Contributors

LS, EH, AB, KS and ML conceived, designed and obtained funding for the study. EH and LS are co-principal investigators. All authors contributed to the statistical analysis plan and DJJF and ML did the statistical analysis. BJ, CRM and DW are members of the study advisory group. LS wrote the first draft and all authors revised the manuscript for intellectual content. The corresponding author attests that all listed authors meet authorship criteria and that no others meeting the criteria have been omitted. LS is guarantor.

## Transparency declaration

LS (guarantor) affirms that the manuscript is an honest, accurate, and transparent account of the study being reported and that no important aspects of the study have been omitted.

## Copyright

The Corresponding Author has the right to grant on behalf of all authors and does grant on behalf of all authors, a worldwide licence to the Publishers and its licensees in perpetuity, in all forms, formats and media (whether known now or created in the future), to i) publish, reproduce, distribute, display and store the Contribution, ii) translate the Contribution into other languages, create adaptations, reprints, include within collections and create summaries, extracts and/or, abstracts of the Contribution, iii) create any other derivative work(s) based on the Contribution, iv) to exploit all subsidiary rights in the Contribution, v) the inclusion of electronic links from the Contribution to third party material where-ever it may be located; and, vi) licence any third party to do any or all of the above.

## References

1. World Health Organisation. WHO Coronavirus Disease (COVID-19) Dashboard, https://covid19.who.int/ (2021, accessed 20/01/2021).

2. Verdery AM, Smith-Greenaway E, Margolis R, et al. Tracking the reach of COVID-19 kin loss with a bereavement multiplier applied to the United States. Proceedings of the National Academy of Sciences 2020; 117: 17695–17701. DOI: 10.1073/pnas.2007476117.

3. Smith KV, Wild J and Ehlers A. The Masking of Mourning: Social Disconnection After Bereavement and Its Role in Psychological Distress. Clinical Psychological Science 2020; 8: 464–476. DOI: 10.1177/2167702620902748.

4. Kentish-Barnes N, Chaize M, Seegers V, et al. Complicated grief after death of a relative in the intensive care unit. European Respiratory Journal 2015; 45: 1341. DOI: 10.1183/09031936.00160014.

5. Selman LE, Chao D, Sowden R, et al. Bereavement support on the frontline of COVID-19: Recommendations for hospital clinicians. Journal of Pain and Symptom Management 2020; 60: e81–e86. DOI: https://doi.org/10.1016/j.jpainsymman.2020.04.024.

6. Probst DR, Gustin JL, Goodman LF, et al. ICU versus Non-ICU hospital death: family member complicated grief, posttraumatic stress, and depressive symptoms. J Palliat Med 2016; 19: 387–393. DOI: 10.1089/jpm.2015.0120.

7. Lobb EA, Kristjanson LJ, Aoun SM, et al. Predictors of complicated grief: A systematic review of empirical studies. Death Stud 2010; 34: 673–698. DOI: 10.1080/07481187.2010.496686.

8. Molina N, Viola M, Rogers M, et al. Suicidal ideation in bereavement: a systematic review. Behavioral Sciences 2019; 9: 53.

9. Burrell A and Selman LE. How do Funeral Practices impact Bereaved Relatives’ Mental Health, Grief and Bereavement? A Mixed Methods Review with Implications for COVID-19. OMEGA-Journal of Death and Dying 2020: 0030222820941296.

10. Mayland CR, Hughes R, Lane S, et al. Are public health measures and individualised care compatible in the face of a pandemic? A national observational study of bereaved relatives’ experiences during the COVID-19 pandemic. Palliative Medicine 2021; 0: 02692163211019885. DOI: 10.1177/02692163211019885.

11. Hanna JR, Rapa E, Dalton LJ, et al. A qualitative study of bereaved relatives’ end of life experiences during the COVID-19 pandemic. Palliative Medicine 2021; 35: 843–851. DOI: 10.1177/02692163211004210.

12. Harrop E, Goss S, Farnell D, et al. Support needs and barriers to accessing support: Baseline results of a mixed-methods national survey of people bereaved during the COVID-19 pandemic. Palliative Medicine 2021; In Press.

13. Covid Bereavement. Grief experience and support needs of people bereaved during the COVID-19 pandemic, https://www.covidbereavement.com/ (2020, accessed 14/01/2021 2021).

14. Claessen SJJ, Francke AL, Sixma HJ, et al. Measuring Relatives’ Perspectives on the Quality of Palliative Care: The Consumer Quality Index Palliative Care. Journal of Pain and Symptom Management 2013; 45: 875–884. DOI: https://doi.org/10.1016/j.jpainsymman.2012.05.007.

15. Harrop E, Morgan F, Longo M, et al. The impacts and effectiveness of support for people bereaved through advanced illness: A systematic review and thematic synthesis. Palliative Medicine 2020; 34: 871–888. DOI: 10.1177/0269216320920533.

16. Harrop E, Scott H, Sivell S, et al. Coping and wellbeing in bereavement: two core outcomes for evaluating bereavement support in palliative care. BMC Palliative Care 2020; 19: 29. DOI: 10.1186/s12904-020-0532-4.

17. Sue Ryder. A Better Grief 2019. London, UK: Sue Ryder.

18. Lobb EA, Kristjanson LJ, Aoun SM, et al. Predictors of complicated grief: a systematic review of empirical studies. Death Stud 2010; 34: 673–698. 2010/09/01. DOI: 10.1080/07481187.2010.496686.

19. Yamaguchi T, Maeda I, Hatano Y, et al. Effects of End-of-Life Discussions on the Mental Health of Bereaved Family Members and Quality of Patient Death and Care. Journal of Pain and Symptom Management 2017; 54: 17–26.e11. DOI: https://doi.org/10.1016/j.jpainsymman.2017.03.008.

20. Miyashita M, Morita T, Sato K, et al. Factors contributing to evaluation of a good death from the bereaved family member’s perspective. Psychooncology 2008; 17: 612–620. 2007/11/10. DOI: 10.1002/pon.1283.

21. Haugen DF, Hufthammer KO, Gerlach C, et al. Good Quality Care for Cancer Patients Dying in Hospitals, but Information Needs Unmet: Bereaved Relatives’ Survey within Seven Countries. Oncologist 2021; 26: e1273–e1284. 2021/06/02. DOI: 10.1002/onco.13837.

22. Eysenbach G. Improving the quality of Web surveys: the Checklist for Reporting Results of Internet E-Surveys (CHERRIES). Journal of medical Internet research 2004; 6: e34–e34. DOI: 10.2196/jmir.6.3.e34.

23. Selman LE, Sowden R and Borgstrom E. ‘Saying goodbye’ during the COVID-19 pandemic: A document analysis of online newspapers with implications for end of life care. Palliative Medicine 2021; 0: 02692163211017023. DOI: 10.1177/02692163211017023.

24. Selman LE, Chamberlain C, Sowden R, et al. Sadness, despair and anger when a patient dies alone from COVID-19: A thematic content analysis of Twitter data from bereaved family members and friends. Palliative Medicine 2021; 0: 02692163211017026. DOI: 10.1177/02692163211017026.

25. Grande GE, Farquhar MC, Barclay SI, et al. Caregiver bereavement outcome: relationship with hospice at home, satisfaction with care, and home death. J Palliat Care 2004; 20: 69–77. 2004/08/31.

26. Hales S, Chiu A, Husain A, et al. The quality of dying and death in cancer and its relationship to palliative care and place of death. J Pain Symptom Manage 2014; 48: 839–851. 2014/04/08. DOI: 10.1016/j.jpainsymman.2013.12.240.

27. Kinoshita H, Maeda I, Morita T, et al. Place of death and the differences in patient quality of death and dying and caregiver burden. J Clin Oncol 2015; 33: 357–363. 2014/12/24. DOI: 10.1200/jco.2014.55.7355.

28. Lei L, Gerlach LB, Powell VD, et al. Caregiver support and place of death among older adults. Journal of the American Geriatrics Society 2021; 69: 1221–1230. DOI: https://doi.org/10.1111/jgs.17055.

29. Bone AE, Finucane AM, Leniz J, et al. Changing patterns of mortality during the COVID-19 pandemic: Population-based modelling to understand palliative care implications. Palliative Medicine 2020; 34: 1193–1201. DOI: 10.1177/0269216320944810.

30. Higginson IJ, Brooks D and Barclay S. Dying at home during the pandemic. BMJ 2021; 373: n1437. DOI: 10.1136/bmj.n1437.

31. Sleeman K, Murtagh F, Kumar R, et al. Better End of Life Care 2021. Dying, death and bereavement during Covid-19. Research Report.. 2021. London, UK.

32. Mitchell S, Oliver P, Gardiner C, et al. Community end-of-life care during the COVID-19 pandemic: findings of a UK primary care survey. BJGP Open 2021; 5: BJGPO.2021.0095. DOI: 10.3399/bjgpo.2021.0095.

33. Eisma MC, Tamminga A, Smid GE, et al. Acute grief after deaths due to COVID-19, natural causes and unnatural causes: An empirical comparison. Journal of Affective Disorders 2021; 278: 54–56. DOI: https://doi.org/10.1016/j.jad.2020.09.049.

34. Breen LJ, Lee SA and Neimeyer RA. Psychological Risk Factors of Functional Impairment After COVID-19 Deaths. J Pain Symptom Manage 2021; 61: e1–e4. 2021/01/22. DOI: 10.1016/j.jpainsymman.2021.01.006.

35. Tang S and Xiang Z. Who suffered most after deaths due to COVID-19? Prevalence and correlates of prolonged grief disorder in COVID-19 related bereaved adults. Globalization and Health 2021; 17: 19. DOI: 10.1186/s12992-021-00669-5.

36. Oluyase A, Hocaoglu M, Cripps R, et al. The challenges of caring for people dying from COVID-19: a multinational, observational study of palliative and hospice services (CovPall). Journal of Pain and Symptom Management 2020 05.02.2021. DOI: https://doi.org/10.1016/j.jpainsymman.2021.01.138.

37. Joaquim RM, Pinto ALCB, Guatimosim RF, et al. Bereavement and psychological distress during COVID-19 pandemics: The impact of death experience on mental health. Current Research in Behavioral Sciences 2021; 2: 100019. DOI: https://doi.org/10.1016/j.crbeha.2021.100019.

38. Mayland CR, Powell R, Clarke G, et al. Bereavement care for ethnic minority communities: A systematic review of access to, models of, outcomes from, and satisfaction with, service provision. PLoS One 2021; 30;16(6):e0252188. DOI: 10.1371/journal.pone.0252188.

39. Ham L, Fransen HP, van den Borne B, et al. Bereaved relatives’ quality of life before and during the COVID-19 pandemic: Results of the prospective, multicenter, observational eQuiPe study. Palliative Medicine; 0: 02692163211034120. DOI: 10.1177/02692163211034120.

40. Eisma MC and Boelen PA. Commentary on: A Call to Action: Facing the Shadow Pandemic of Complicated Forms of Grief. OMEGA - Journal of Death and Dying; 0: 00302228211016227. DOI: 10.1177/00302228211016227.

41. Lee SA and Neimeyer RA. Pandemic Grief Scale: A screening tool for dysfunctional grief due to a COVID-19 loss. Death Studies 2020: 1–11. DOI: 10.1080/07481187.2020.1853885.

