## Supplementary file 1 for "Place, cause and expectedness of death and relationship to the deceased are associated with poorer experiences of end-of-life care and challenges in early bereavement: Risk factors from an online survey of people bereaved during the COVID-19 pandemic"

### The grief experiences and support needs of people bereaved during the COVID-19 pandemic

#### Page 1

##### We invite you to take part in a study

Thank you for taking the time to read about this study.

We recognise what a difficult time this may be for you at the moment and we would like to express our sincere condolences for your loss.

**Background and purpose of the study**

We are doing this research to understand the grief experiences and support needs of people who have been bereaved either due to COVID-19 or another cause of death during the pandemic.

We are interested in your experiences at the end of life of your loved one (family or close friends). Also, your experiences of grieving, any support that you feel you have needed during your bereavement and what support you have received.

We hope that your responses to our questions will help us to identify ways to improve end of life care during the pandemic and the support that is available for people who are bereaved. We will also provide information about services that could support you during this time.

This survey is being carried out by Cardiff University and the University of Bristol and is funded by the Economic and Social Research Council. It should take around 20-30 minutes to complete, depending on how much you would like to share about your experiences.

**Who can take part:**

- Anyone aged 18 or over
- Anyone who has experienced the loss of a close family member, friend or other loved one who died in the UK since the 16th March 2020, *whether this was caused by COVID-19 or another cause of death.*

At the end of the survey, you will be asked if you would like to be contacted to discuss some of your experiences in more depth in an interview with a researcher. We will also ask your permission to send you two similar questionnaires in the next year to see how things may have changed for you. If you agree to this, we will send these out to you around 7 and 13 months after the date that your loved one died.

For more information, please read the following sections.

**Risks and benefits:**

This survey asks questions about the loss of your loved one(s) during the COVID-19 pandemic and could become difficult to answer. Should you become upset and feel you need support during or after you have completed the survey, please get in touch with our researcher Dr Emily Harrop who can guide you towards appropriate services. Alternatively, you can access support from these services:

- Marie Curie Bereavement Support: 0800 090 2309 https://www.mariecurie.org.uk/help/support/bereaved-family-friends/dealing-grief/bereavement-or-grief-counselling
- Cruse Bereavement Care: 0808 808 1677
- NHS Bereavement Helpline: 0800 2600 400 https://www.nhs.uk/conditions/stress-anxiety-depression/coping-with-bereavement/
- The Good Grief Trust: https://www.thegoodgrieftrust.org/

When using the internet, there can be a risk of compromising privacy, confidentiality and/or anonymity. We are using a secure online survey platform to minimise this risk (Jisc Online Surveys), further information is provided below.

By completing this survey on your experiences of bereavement during COVID-19, you are helping us to learn about the key issues that you have faced and to identify ways of improving the care provided at the end of life and during bereavement.

**Voluntary participation:**

Participation in this study is completely voluntary. You may choose not to answer some or any of the questions. If you decide to withdraw at any stage, you do not have to give any reason and your decision will be respected.

If you would like to withdraw your data from the study, please contact the researcher using the contact details below. However, please be aware that it may not be possible to withdraw data that has been anonymised and analysed.

**Confidentiality:**

Your answers will be seen only by the research team based at Cardiff University. Ethical approval to conduct the study has been granted by Cardiff University School of Medicine Research Ethics Committee. Any information that you provide, including any quotations that we use from your responses, will be anonymised to make sure you cannot be indirectly identified.

The information you provide will remain confidential, however if your answers were to identify any area of sub-standard care, malpractice or abuse, the researcher has a statutory duty to raise this with the appropriate bodies.

**Data protection and usage:**

By participating in this survey, you are agreeing that we may include your survey responses (anonymously) in future reports, professional journals and future research. Some anonymised data will also be shared with colleagues in Europe, to enable international comparisons to be made. Anonymised study findings may be presented at professional conferences, in promotional material (e.g. press releases) and in educational settings.

In line with Cardiff University policies and General Data Protection Regulations (GDPR; EU 2016/679) and the Data Protection Act 2018 (DPA 2018), your research data will be kept securely for 10 years after the study is completed and securely destroyed at the end of the 10 years. If you provide us with contact details for follow up questionnaires, interviews or to be sent further information about this or other research studies, these details will be kept securely for up to a year after the study ends and securely destroyed once they are no longer needed. Anonymised research data will also be archived so that it can be used by other researchers in the future.

All data collected in the on-line survey will be held securely by the survey software provider Jisc Online Survey (www.jisc.ac.uk) under contract and then retained by Cardiff University in accordance with the GDPR; EU 2016/679 and DPA 2018. For more information on data protection, please follow the link: General Data Protection Regulation (GDPR; EU 2016/679). Cookies (personal data stored by your web browser) are not used in this survey.

**Instructions for completion**

- This survey contains 4 sections and 36 questions.
- It should take around 20- 30 minutes to complete.
- The survey can be completed in more than one sitting by clicking the 'finish later' link at the bottom of the page. You will then need to either bookmark your unique URL link in your browser or ask for it to be emailed to you. You can navigate the survey and edit your answers up to the point you click the 'finish survey' button.

If you would like a paper or Microsoft Word version of the survey or a copy of the survey in Welsh or another community language please contact us using the contact details provided. Please also contact us if you would prefer to complete the survey over the phone or by Zoom with one of our researchers instead.

**Contact information:** If you have any concerns or questions relating to this research project, please email the researcher Dr Emily Harrop, or TEL: 02920 687184. Additionally, if you have a complaint or other queries, contact the research centre's Director Professor Annmarie Nelson on.

Alternatively, you can contact the research centre via 02920 687175 or to be directed to the appropriate individual. Our postal address is: Marie Curie Palliative Care Research Centre, Cardiff University, 8th Floor Neuadd Meirionnydd, Heath Park, Cardiff, CF14 4YS.

### Page 2

**Many thanks for reading about and deciding to take part in this survey**

**Consent**

By participating in this survey, you agree that you have read and understood the information provided above and that you are aged 18 or over.

I confirm that I have read and understood the information provided about the purpose of this study and how my data will be used. I agree to take part in the following survey knowing that all questions are optional and I can finish the survey at any point.

Please tick the box to agree with the above statement: ✱ *Required*

- [ ] I agree

**Thank you for your help, you will now be able to start completing the survey.**

### Page 3

#### Part A: Information about you and your bereavement

**This section collects information about your family member(s) or close friend(s) who you lost during the COVID 19 pandemic (since 16th March 2020).**

**A1. Who was it that died?** If you have lost more than one person since the start of the pandemic (16th March 2020), please select all answers that apply:

- ☐ My husband or male partner
- ☐ My wife or female partner
- ☐ My mother
- ☐ My father
- ☐ My brother
- ☐ My sister
- ☐ My son
- ☐ My daughter
- ☐ My grandparent
- ☐ My aunt or uncle
- ☐ My grandchild
- ☐ My friend
- ☐ My colleague
- ☐ Other

If you selected Other, please specify:

**If you have lost more than one person, please choose one of these people to write about when answering these next sets of questions. Please tell us here who you will be referring to and why you have chosen this person:**

**A2. How old were they?**

**A3. Where did they die?**

- In hospital
- In their home
- In a hospice
- In a care home
- I don't know
- Other

If you selected Other, please specify:

**A4. Please tell us when they died (if possible please provide the date)**

**A5. Do you know the cause of death?**

- Confirmed COVID-19 infection
- Suspected COVID-19 infection
- Cancer
- Don't know
- Other

If you selected Other, please specify:

**A6. Did you expect your loved one to die around this time? (e.g. if they had a terminal illness)**

- Yes
- No
- Don't know

If you would like to comment on your answer to this question please use this text box:

### Page 4

#### Part B. Your experiences at the end of life of your loved one

In this section, we would like to find out more about your experiences just before and after the death of your loved one (e.g. in hospital, hospice or when caring at home).

We would like to know how you feel these experiences have affected you and if better support could have been provided.

*If you have lost more than one person, please continue to tell us about the same person that you discussed in Section A.*

#### Communication with healthcare/care professionals before and immediately after the death of your loved one

**B1. Did the care professionals involve you in decisions about the care for your sick loved one?**

- Never
- Sometimes
- Usually
- Always
- Not relevant to my situation (e.g. not next of kin, because none were involved)

**B2. Did you know the contact details for the professional responsible for their care?**

- Yes
- No
- Not sure
- Not relevant to my situation

**B3. Did you receive information about the approaching death?**

- No, not at all
- A bit of information
- Yes, I was fully informed
- Not relevant to my situation

If yes, who gave you this information?

**B4. Did you feel well supported by the healthcare professionals immediately after the death of your loved one?**

- Very well supported
- Fairly well supported
- A little bit supported
- Not at all supported
- Not relevant to my situation (e.g. because none were involved or not next of kin)

**B5. Were you contacted again by the hospital or care provider following their death?**

- Yes
- No
- Not relevant to my situation

If yes, approximately how long after the death did they contact you?

Who was it that contacted you?

**B6. Did they provide information about bereavement support services? (tick all that apply)**

- ☐ Yes (at the time of death)
- ☐ Yes (during follow up call)
- ☐ No
- ☐ Not relevant to my situation

**B7. Please indicate how these communications with healthcare/care professionals before and after the death of your loved one took place. Please also rate how effective you found the method of communication (tick all that apply):**

| | Effective | Slightly effective | Not effective | Not used |
|---|---|---|---|---|
| By phone | ☐ | ☐ | ☐ | ☐ |
| By video call (e.g. Zoom, Skype) | ☐ | ☐ | ☐ | ☐ |
| In person (face to face) | ☐ | ☐ | ☐ | ☐ |
| In writing (e.g. letter, leaflet, e-mail) | ☐ | ☐ | ☐ | ☐ |

Please tell us about any communications that worked well or not so well, and any improvements that could be made to the communications you received from professionals at this time. We will use this information to help improve services.

**B8. When your loved one died, did you experience any of the following? (Please tick all that apply):**

- ☐ Unable to visit them prior to their death
- ☐ Limited contact with them in last days of their life
- ☐ Unable to say goodbye as I would have liked
- ☐ Restricted funeral arrangements
- ☐ Social isolation and loneliness
- ☐ Limited contact with other close relatives or friends

If you would like to write about these or any other experiences around the time that your loved one died, please use the text box below. For example, you may like to tell us about how these experiences (positive or negative) have affected you or how you think the care provided at this time could have been better:

**B9. To help us better understand how you have been adjusting to your loss. Please indicate your response to the following attitudes:**

More info

| | Strongly agree | Agree | Neither agree nor disagree | Disagree | Strongly disagree |
|---|---|---|---|---|---|
| 1. I feel able to face the pain which comes with loss. | ☐ | ☐ | ☐ | ☐ | ☐ |
| 2. For me, it is difficult to switch off thoughts about the person I have lost. | ☐ | ☐ | ☐ | ☐ | ☐ |
| 3. I feel very aware of my inner strength when faced with grief. | ☐ | ☐ | ☐ | ☐ | ☐ |
| 4. I believe that I must be brave in the face of loss. | ☐ | ☐ | ☐ | ☐ | ☐ |
| 5. I feel that I will always carry the pain of grief with me. | ☐ | ☐ | ☐ | ☐ | ☐ |
| 6. For me, it is important to keep my grief under control. | ☐ | ☐ | ☐ | ☐ | ☐ |
| 7. Life has less meaning for me after this loss. | ☐ | ☐ | ☐ | ☐ | ☐ |
| 8. I think it’s best just to get on with life in spite of this loss. | ☐ | ☐ | ☐ | ☐ | ☐ |

| | | | | | |
|---|---|---|---|---|---|
| 9. It may not always feel like it but I do believe that I will come through this experience of grief. | ☐ | ☐ | ☐ | ☐ | ☐ |

### Page 5

#### Part C. Bereavement support

**This next section includes questions on the types of support you might have been using to help you cope since your loss and any needs for support that you have been experiencing.**

**C1. What types of support have you used to cope with your loss? Please select all that apply:**

- ☐ Support by family or friends
- ☐ GP or other member of staff at the GP surgery
- ☐ Telephone helpline support (e.g. bereavement helpline)
- ☐ Online community support via written comments (e.g. Facebook group, online chat forum)
- ☐ Informal support group (e.g. social group for bereaved people)
- ☐ Bereavement support group (e.g. group discussions about bereavement guided by a facilitator; or group counselling)
- ☐ One-to-one support (e.g. individual counselling)
- ☐ Specialist mental health support
- ☐ Other
- ☐ No support used

If you selected Other, please specify:

Please tell us who has provided this support and how it was provided (e.g. face to face, by telephone, Zoom etc):

Please tell us how helpful you have found this support. If possible, please also tell us some of the ways in which you have been helped by the support:

**C2. Have you experienced difficulties getting support and help following your bereavement?**

| | Yes | Somewhat | No | I've not tried to get their support |
|---|---|---|---|---|
| From friends and family | ☐ | ☐ | ☐ | ☐ |
| From GP surgery | ☐ | ☐ | ☐ | ☐ |
| From bereavement services | ☐ | ☐ | ☐ | ☐ |

**C3. Do any of the responses below describe your experiences? Please select all that apply:**

- ☐ I have not wanted any support from bereavement services because my family and friends provide me with enough support
- ☐ I have not wanted any support from bereavement services because I do not think it would help me
- ☐ I do not know how to get support from bereavement services
- ☐ I have felt uncomfortable asking for support from bereavement services
- ☐ I have felt uncomfortable asking for help or support from friends or family
- ☐ The support I wanted from bereavement services was not available to me
- ☐ Friends or family have not been able to support me in the way I wanted

**C4. If relevant, please briefly describe any difficulties you faced getting support from friends, family or bereavement services:**

**C5. Over the last three months please tell us how much support or help you feel that you have needed with the following:**

| | High level of support needed | Fairly high level of support needed | Moderate level of support needed | Little support needed | No support needed |
|---|---|---|---|---|---|
| Practical tasks e.g. managing the funeral, registering the death, other paperwork etc. | ☐ | ☐ | ☐ | ☐ | ☐ |
| Getting relevant information and advice e.g. legal, financial, available support | ☐ | ☐ | ☐ | ☐ | ☐ |
| Looking after myself/family e.g. getting food, medication, childcare | ☐ | ☐ | ☐ | ☐ | ☐ |
| Dealing with my feelings about being without my loved one | ☐ | ☐ | ☐ | ☐ | ☐ |
| Dealing with my feelings about the way my loved one died | ☐ | ☐ | ☐ | ☐ | ☐ |
| Expressing my feelings and feeling understood by others | ☐ | ☐ | ☐ | ☐ | ☐ |
| Feeling comforted and reassured | ☐ | ☐ | ☐ | ☐ | ☐ |
| Loneliness and social isolation | ☐ | ☐ | ☐ | ☐ | ☐ |

| | | | | | |
|---|---|---|---|---|---|
| Managing and maintaining my relationships with friends and family | ☐ | ☐ | ☐ | ☐ | ☐ |
| Finding balance between grieving and other areas of life | ☐ | ☐ | ☐ | ☐ | ☐ |
| Participating in work, leisure or other regular activities (e.g. shopping, housework) | ☐ | ☐ | ☐ | ☐ | ☐ |
| Feelings of anxiety and depression | ☐ | ☐ | ☐ | ☐ | ☐ |
| Regaining sense of purpose and meaning in life | ☐ | ☐ | ☐ | ☐ | ☐ |

Please provide further details on these or any other areas of support that you feel you need now or may have needed earlier on in your bereavement:

**C6. If you would like to make any additional comments, including anything else you have found particularly helpful or difficult at this time (and have not already discussed) please write them below:**

### Page 6

**PART D:**

**In this last section we would like to ask you a few additional questions to help us understand who answered our questionnaire (for instance different age groups). You will never be identified as we will combine the answers from all the respondents.**

**D1. Age:** How old are you?

**D2. Highest qualification:** What is the highest level of formal education that you have completed? (*Please tick one only*)

- No qualifications
- GCSEs/ O Levels/ CSEs
- A Levels/ GNVQs/BTEC
- Trade apprenticeship
- ONC/OND/City & Guilds
- HNC/HND
- University First Degree (e.g. BA, BSc)
- Postgraduate Degree (e.g. MA, MSc, PhD)
- Postgraduate Qualification (e.g. certificate or diploma)
- Other

If you selected Other, please specify:

**D3. Occupation:** Which of these best describes what you are doing at present? (*If more than one of these applies to you, please tick the main ONE*)

- Full-time paid work (30 hours or more per week)
- Part-time paid work (Under 30 hours per week)
- Full time education at school, college or university
- Unemployed
- Permanently sick/disabled
- Fully retired from work
- Looking after the home
- Caring for a family member or loved one
- Other/Doing something else

If you selected Other, please specify:

**D4. Have you become unemployed since the start of the Covid-19 Pandemic (16th March 2020)?**

- Yes
- No

**D5. What is your current or most recent job?**

**D6. Your household:** How many people live in your house (including yourself)?

**D7. Your neighbourhood:** What is your full UK postcode? If you are currently living outside the UK please state your country of residence.

**D8. Ethnicity:** Tick **one** box below which best describes your ethnic group.

White:

- British
- English
- Scottish
- Welsh
- Irish
- Northern Irish
- Any other white background

If you selected Other, please specify:

Mixed:

- White and Black Caribbean
- White and Black African
- White and Asian
- Any other mixed/multiple ethnic background

If you selected Other, please specify:

Asian or Asian British:

- Indian
- Pakistani
- Bangladeshi
- Chinese
- Any other Asian background

If you selected Other, please specify:

Black/African/Caribbean/ Black British:

- African
- Caribbean
- Any other Black/African/ Caribbean background

If you selected Other, please specify:

Other ethnic group:

- Arab
- Any other ethnic group

If you selected Other, please specify:

**D9. Do you have any religious or spiritual beliefs?**

- Yes
- No
- Prefer not to say

If yes, please select one of the following options:

- Agnostic
- Buddhism
- Christianity (all denominations)
- Hinduism
- Islam
- Judaism
- Sikhism

- Spiritual but not religious
- Other

If you selected Other, please specify:

**D10. Gender Identity:** Previous research has indicated that people's gender identity and sexual orientation can affect their use of bereavement support. This is why we are asking participants to share this information, if willing to do so. Please describe your gender identity:

- Male
- Female
- Non-binary
- Prefer not to say
- Other

If you selected Other, please specify:

**Is this the same sex which you were assigned at birth?**

- Yes
- No
- Prefer not to say

**D11. Sexual orientation:** Please describe your sexual identity (e.g.

heterosexual/straight, gay/lesbian etc.)

If you prefer not to say please tick the box below:

- Prefer not to say

**D12. Your health**: This information is to enable us to consider how health issues might affect/be affected by bereavement. Do you have any particular medical condition(s)?

- Yes
- No

If yes, please tell us what condition(s) you have and any treatments you are receiving:

**D13. GP visits**: Can you remember roughly how many times you have visited the GP in the **last three months**?

**D14. Previous bereavements:** Have you experienced the loss of any other close family members, friends or loved ones in the 12 months before the start of the pandemic (16th March 2020)?

- Yes
- No

**D15. Finally, please tell us where you found out about this survey?**

### Page 7

**Many thanks for completing this survey.**

Your responses will help to enable others who are bereaved to access the support they need. We appreciate that this may have been difficult and painful for you and we are very grateful for your contribution.

**Invitation to take part:** We would like to send you *two similar questionnaires* over the coming months to see how things may have changed for you. If you are happy for us to do so, please indicate below and fill in your name and contact details at the end of this section:

- Yes
- No

We would also like to invite 20-30 people to talk about their experiences in more depth in an *informal interview* carried out over the phone or by Zoom. If you would be happy to be contacted with more information about taking part in an interview please indicate below and leave your name and contact details at the end of this section:

- Yes
- No

If you would like us to send you information about other *similar research studies* that you may be interested in taking part in, please indicate below and leave your name and contact details at the end of this section:

- Yes
- No

If you would like to receive *updates about the study*, including a copy of the results, please indicate below and leave your name and contact details at the end of this section:

- ○ Yes
- ○ No

**Contact Details:**

If you answered yes to receiving further communications from us (as above), please tell us your name and how would you like to receive this information. Please be assured that this information will be strictly confidential and not shared with anyone else:

- ○ By email
- ○ By post

Name:

My email address is:

My address is:

#### Page 8: Final page

***Thank you again for your help***

**We are extremely grateful for your contribution.**

**Here are our contact details should you wish to get in touch: Emily Harrop or 02920687184**

**If you would like to talk to someone about your bereavement, you can access support from these services:**

- Marie Curie Bereavement Support: 0800 090 2309 https://www.mariecurie.org.uk/help/support/bereaved-family-friends/dealing-grief/bereavement-or-grief-counselling
- Cruse Bereavement Care: 0808 808 1677
- NHS Bereavement Helpline: 0800 2600 400 https://www.nhs.uk/conditions/stress-anxiety-depression/coping-with-bereavement/
- The Good Grief Trust: https://www.thegoodgrieftrust.org/
