## Supplementary file 3 Tables for "Place, cause and expectedness of death and relationship to the deceased are associated with poorer experiences of end-of-life care and challenges in early bereavement: Risk factors from an online survey of people bereaved during the COVID-19 pandemic"

|  | | *n* | Never | Sometimes | Usually | Always | Mean | Median | SD | *h* (Effect Size),  *P* |
| --- | --- | --- | --- | --- | --- | --- | --- | --- | --- | --- |
| Highest qualification | None / GCSEs | 96 | 37.5% | 24.0% | 12.5% | 26.0% | 2.27 | 2.39 | 2.51 | 0.29 (Small)  0.183 |
|  | A-level etc. | 115 | 27.8% | 28.7% | 20.0% | 23.5% | 2.00 | 2.00 | 2.00 |  |
|  | HND / Degree etc. | 356 | 23.9% | 29.8% | 17.7% | 28.7% | 1.22 | 1.13 | 1.14 |  |
| Where did they die? | Hospital | 350 | 35.4% | 29.1% | 15.4% | 20.0% | 2.20 | 2.00 | 1.13 | 0.88 (Large)  <0.001* |
|  | Home | 100 | 13.0% | 24.0% | 19.0% | 44.0% | 2.94 | 3.00 | 1.10 |  |
|  | Hospice | 35 | 8.6% | 22.9% | 22.9% | 45.7% | 3.06 | 3.00 | 1.03 |  |
|  | Care home | 78 | 17.9% | 34.6% | 19.2% | 28.2% | 2.58 | 2.00 | 1.09 |  |
|  | Other / Don’t Know | 5 | 20.0% | 20.0% | 0.0% | 60.0% | 3.00 | 4.00 | 1.41 |  |
| Cause of death | COVID (Confirmed or Suspected) | 266 | 39.1% | 29.7% | 13.9% | 17.3% | 2.09 | 2.00 | 1.10 | 0.47 (Medium) <0.001* |
|  | Non-Covid | 303 | 16.8% | 27.4% | 20.1% | 35.6% | 2.75 | 3.00 | 1.12 |  |
| Gender Identity | Male | 60 | 18.3% | 30.0% | 21.7% | 30.0% | 2.63 | 3.00 | 1.10 | 0.1 (Small)  0.089 |
|  | Female | 506 | 28.3% | 27.9% | 16.8% | 27.1% | 2.43 | 2.00 | 1.16 |  |
|  | Other | 3 | 0.0% | 100.0% | 0.0% | 0.0% | 2.45 | 2.00 | 1.16 |  |
| Ethnicity | Non-BAME | 541 | 26.8% | 28.7% | 17.2% | 27.4% | 2.41 | 2.00 | 1.19 | 0.06 (Small)  0.979 |
|  | BAME | 27 | 29.6% | 25.9% | 18.5% | 25.9% | 2.53 | 2.00 | 1.18 |  |
| Who was it that died? | Partner | 140 | 25.7% | 27.1% | 15.7% | 31.4% | 2.53 | 2.00 | 1.18 | 0.60 (Medium)  0.154 |
|  | Parent | 356 | 25.6% | 29.5% | 17.7% | 27.2% | 2.47 | 2.00 | 1.14 |  |
|  | Grandparent | 25 | 32.0% | 28.0% | 16.0% | 24.0% | 2.32 | 2.00 | 1.18 |  |
|  | Sibling | 15 | 46.7% | 20.0% | 20.0% | 13.3% | 2.00 | 2.00 | 1.13 |  |
|  | Child | 9 | 22.2% | 11.1% | 33.3% | 33.3% | 2.78 | 3.00 | 1.20 |  |
|  | Other family member | 20 | 30.0% | 40.0% | 15.0% | 15.0% | 2.15 | 2.00 | 1.04 |  |
|  | Colleague or friend | 5 | ─ | ─ | ─ | ─ |  |  |  |  |
| Religious beliefs | Yes | 280 | 26.4% | 25.7% | 17.9% | 30.0% | 2.51 | 2.00 | 1.18 | 0.11 (Small)  0.463 |
|  | No | 239 | 26.4% | 31.0% | 18.0% | 24.7% | 2.41 | 2.00 | 1.13 |  |
| Was the death expected? | Yes | 94 | 5.3% | 22.3% | 26.6% | 45.7% | 3.13 | 3.00 | 0.94 | 0.56 (Medium) <0.001* |
|  | No | 439 | 32.6% | 30.1% | 15.0% | 22.3% | 2.27 | 2.00 | 1.14 |  |
| Region of the UK | Northern Ireland | 20 | 20.0% | 25.0% | 10.0% | 45.0% | 2.8 | 3 | 1.24 | 0.69 (Medium / Large),  0.268 |
|  | Wales | 50 | 22.0% | 34.0% | 16.0% | 28.0% | 2.5 | 2 | 1.129 |  |
|  | West Midlands | 45 | 33.3% | 31.1% | 11.1% | 24.4% | 2.27 | 2 | 1.176 |  |
|  | North East | 30 | 26.7% | 26.7% | 10.0% | 36.7% | 2.57 | 2 | 1.251 |  |
|  | Greater London | 49 | 24.5% | 18.4% | 26.5% | 30.6% | 2.63 | 3 | 1.167 |  |
|  | South East | 62 | 30.6% | 27.4% | 27.4% | 14.5% | 2.26 | 2 | 1.055 |  |
|  | North West | 80 | 25.0% | 31.3% | 11.3% | 32.5% | 2.51 | 2 | 1.191 |  |
|  | East Midlands | 35 | 40.0% | 25.7% | 14.3% | 20.0% | 2.14 | 2 | 1.167 |  |
|  | South West | 42 | 19.0% | 31.0% | 16.7% | 33.3% | 2.64 | 2.5 | 1.144 |  |
|  | Scotland | 41 | 22.0% | 26.8% | 12.2% | 39.0% | 2.68 | 3 | 1.213 |  |
|  | East of England | 33 | 27.3% | 33.3% | 27.3% | 12.1% | 2.24 | 2 | 1.001 |  |
|  | Yorkshire & the Humber | 41 | 22.0% | 31.7% | 12.2% | 34.1% | 2.59 | 2 | 1.183 |  |
| IMD  Decile (England only) | 1 | 23 | 43.5% | 21.7% | 8.7% | 26.1% | 2.17 | 2 | 1.267 | 0.50 (Medium),  0.552 |
|  | 2 | 35 | 25.7% | 25.7% | 14.3% | 34.3% | 2.57 | 2 | 1.22 |  |
|  | 3 | 41 | 26.8% | 24.4% | 12.2% | 36.6% | 2.59 | 2 | 1.245 |  |
|  | 4 | 45 | 20.0% | 40.0% | 15.6% | 24.4% | 2.44 | 2 | 1.078 |  |
|  | 5 | 50 | 24.0% | 30.0% | 12.0% | 34.0% | 2.56 | 2 | 1.198 |  |
|  | 6 | 43 | 30.2% | 25.6% | 11.6% | 32.6% | 2.47 | 2 | 1.241 |  |
|  | 7 | 47 | 25.5% | 31.9% | 17.0% | 25.5% | 2.43 | 2 | 1.137 |  |
|  | 8 | 43 | 25.6% | 25.6% | 30.2% | 18.6% | 2.42 | 2 | 1.074 |  |
|  | 9 | 36 | 27.8% | 25.0% | 25.0% | 22.2% | 2.42 | 2 | 1.131 |  |
|  | 10 | 43 | 27.9% | 32.6% | 25.6% | 14.0% | 2.26 | 2 | 1.131 |  |

* *P* < 0.05

**Table S1:** Descriptive statistics (row percentages, means, and medians) for question “Did the care professionals involve you in decisions about the care for your sick loved one?”. *P*-values are from chi-squared analysis of a contingency table. Effect sizes are estimated from Cohen’s *h*, where *h* = 0.2: small effect, *h* = 0.5: medium effect, *h* ≥ 0.8: large effect.

|  | | *n* | Yes | No | *h* (Effect Size),  *P* |
| --- | --- | --- | --- | --- | --- |
| Highest qualification | None / GCSEs | 85 | 56.5% | 43.5% | 0.30 (Small),  0.186 |
|  | A-level etc. | 107 | 68.2% | 31.8% |  |
|  | HND / Degree etc. | 352 | 65.9% | 34.1% |  |
| Where did they die? | Hospital | 329 | 55.0% | 45.0% | 0.80 (Large),  0.001* |
|  | Home | 101 | 80.2% | 19.8% |  |
|  | Hospice | 34 | 70.6% | 29.4% |  |
|  | Care home | 77 | 83.1% | 16.9% |  |
|  | Other / Don’t Know | 5 | 60.0% | 40.0% |  |
| Cause of death | COVID (Confirmed or Suspected) | 255 | 58.4% | 41.6% | 0.31 (Small),  0.005* |
|  | Non-Covid | 291 | 70.1% | 29.9% |  |
| Gender Identity | Male | 58 | 67.2% | 32.8% | 0.07 (Small),  0.925 |
|  | Female | 482 | 64.5% | 35.5% |  |
|  | Other | 4 | 75.0% | 25.0% |  |
| Ethnicity | Non-BAME | 519 | 64.4% | 35.6% | 0.36 (Small / Medium),  0.213 |
|  | BAME | 26 | 76.9% | 23.1% |  |
| Who was it that died? | Partners | 127 | 74.0% | 26.0% | 1.11 (Large), 0.001* |
|  | Parents | 335 | 65.1% | 34.9% |  |
|  | Grandparents | 27 | 63.0% | 37.0% |  |
|  | Sibling | 16 | 31.3% | 68.8% |  |
|  | Child | 8 | 62.5% | 37.5% |  |
|  | Other family member | 23 | 52.2% | 47.8% |  |
|  | Colleague or friend | 11 | 27.3% | 72.7% |  |
| Religious beliefs | Yes | 266 | 63.9% | 36.1% | 0.14 (Small), 0.253 |
|  | No | 229 | 69.0% | 31.0% |  |
| Was the death expected? | Yes | 94 | 86.2% | 13.8% | 0.83 (Large),  <0.001* |
|  | No | 417 | 58.5% | 41.5% |  |
| Region of the UK | Northern Ireland | 20 | 65.0% | 35.0% | 0.52 (Medium),  0.778 |
|  | Wales | 50 | 60.0% | 40.0% |  |
|  | West Midlands | 41 | 61.0% | 39.0% |  |
|  | North East | 27 | 77.8% | 22.2% |  |
|  | Greater London | 52 | 63.5% | 36.5% |  |
|  | South East | 52 | 63.5% | 36.5% |  |
|  | North West | 82 | 73.2% | 26.8% |  |
|  | East Midlands | 34 | 58.8% | 41.2% |  |
|  | South West | 41 | 73.2% | 26.8% |  |
|  | Scotland | 40 | 70.0% | 30.0% |  |
|  | East of England | 32 | 62.5% | 37.5% |  |
|  | Yorkshire & the Humber | 38 | 65.8% | 34.2% |  |
| IMD  Decile (England only) | 1 | 23 | 65.2% | 34.8% | 0.56 (Medium),  0.861 |
|  | 2 | 34 | 70.6% | 29.4% |  |
|  | 3 | 37 | 67.6% | 32.4% |  |
|  | 4 | 45 | 64.4% | 35.6% |  |
|  | 5 | 50 | 66.0% | 34.0% |  |
|  | 6 | 39 | 74.4% | 25.6% |  |
|  | 7 | 45 | 66.7% | 33.3% |  |
|  | 8 | 48 | 66.7% | 33.3% |  |
|  | 9 | 32 | 53.1% | 46.9% |  |
|  | 10 | 37 | 73.0% | 27.0% |  |

* *P* < 0.05

**Table S2:** Descriptive statistics (row percentages) for the question “Did you know the contact details for the professional responsible for their care?”. *P*-values are from chi-squared analysis of a contingency table. Effect sizes are estimated from Cohen’s *h*, where *h* = 0.2: small effect, *h* = 0.5: medium effect, *h* ≥ 0.8: large effect.

|  | | *n* | No, not at all (=1) | A bit of information (=2) | Yes, fully informed  (=3) | Mean | Median | SD | *h* (Effect Size),  *P* |
| --- | --- | --- | --- | --- | --- | --- | --- | --- | --- |
| Highest qualification | None / GCSEs | 97 | 26.8% | 40.2% | 33.0% | 2.06 | 2.00 | 0.78 | 0.25 (Small),  0.113 |
|  | A-level etc. | 123 | 17.9% | 51.2% | 30.9% | 2.13 | 2.00 | 0.69 |  |
|  | HND / Degree etc. | 404 | 19.1% | 41.3% | 39.6% | 2.21 | 2.00 | 0.74 |  |
| Where did they die? | Hospital | 387 | 20.9% | 44.7% | 34.4% | 2.13 | 2.00 | 0.73 | 0.59 (Medium),  0.22 |
|  | Home | 112 | 22.3% | 42.0% | 35.7% | 2.13 | 2.00 | 0.75 |  |
|  | Hospice | 36 | 11.1% | 30.6% | 58.3% | 2.47 | 3.00 | 0.70 |  |
|  | Care home | 84 | 16.7% | 44.0% | 39.3% | 2.23 | 2.00 | 0.72 |  |
|  | Other / Don’t Know | 5 | 20.0% | 20.0% | 60.0% | 2.40 | 3.00 | 0.89 |  |
| Cause of death | COVID (Confirmed or Suspected) | 295 | 19.0% | 48.8% | 32.2% | 2.13 | 2.00 | 0.70 | 0.24 (Small),  0.018* |
|  | Non-Covid | 330 | 21.2% | 37.9% | 40.9% | 2.20 | 2.00 | 0.76 |  |
| Gender Identity | Male | 69 | 18.8% | 34.8% | 46.4% | 2.28 | 2.00 | 0.77 | 0.24 (Small),  0.072 |
|  | Female | 551 | 20.0% | 44.5% | 35.6% | 2.16 | 2.00 | 0.73 |  |
|  | Other | 5 | 60.0% | 20.0% | 20.0% | 1.60 | 1.00 | 0.89 |  |
| Ethnicity | Non-BAME | 597 | 19.8% | 43.6% | 36.7% | 2.17 | 2.00 | 0.73 | 0.18 (Small),  0.501 |
|  | BAME | 28 | 28.6% | 35.7% | 35.7% | 2.07 | 2.00 | 0.81 |  |
| Who was it that died? | Partners | 132 | 18.9% | 36.4% | 44.7% | 2.26 | 2.00 | 0.76 | 0.95 (Large),  0.096 |
|  | Parents | 375 | 18.4% | 45.3% | 36.3% | 2.18 | 2.00 | 0.72 |  |
|  | Grandparent | 43 | 25.6% | 44.2% | 30.2% | 2.05 | 2.00 | 0.75 |  |
|  | Sibling | 21 | 19.0% | 38.1% | 42.9% | 2.24 | 2.00 | 0.77 |  |
|  | Child | 8 | 12.5% | 75.0% | 12.5% | 2.00 | 2.00 | 0.54 |  |
|  | Other family member | 32 | 28.1% | 46.9% | 25.0% | 1.97 | 2.00 | 0.74 |  |
|  | Colleague or friend | 15 | 46.7% | 26.7% | 26.7% | 1.80 | 2.00 | 0.86 |  |
| Religious beliefs | Yes | 302 | 18.5% | 41.1% | 40.4% | 2.22 | 2.00 | 0.74 | 0.13 (Small),  0.284 |
|  | No | 265 | 22.3% | 43.4% | 34.3% | 2.12 | 2.00 | 0.74 |  |
| Was the death expected? | Yes | 106 | 12.3% | 32.1% | 55.7% | 2.43 | 3.00 | 0.70 | 0.54 (Medium),  <0.001* |
|  | No | 478 | 22.4% | 46.0% | 31.6% | 2.09 | 2.00 | 0.73 |  |
| Region of the UK | Northern Ireland | 22 | 22.7% | 27.3% | 50.0% | 2.27 | 2.5 | 0.827 | 0.67 (Medium / Large),  0.299 |
|  | Wales | 57 | 28.1% | 49.1% | 22.8% | 1.95 | 2 | 0.718 |  |
|  | West Midlands | 47 | 14.9% | 44.7% | 40.4% | 2.26 | 2 | 0.706 |  |
|  | North East | 33 | 18.2% | 54.5% | 27.3% | 2.09 | 2 | 0.678 |  |
|  | Greater London | 63 | 25.4% | 34.9% | 39.7% | 2.14 | 2 | 0.8 |  |
|  | South East | 70 | 17.1% | 52.9% | 30.0% | 2.13 | 2 | 0.679 |  |
|  | North West | 88 | 18.2% | 44.3% | 37.5% | 2.19 | 2 | 0.725 |  |
|  | East Midlands | 36 | 16.7% | 47.2% | 36.1% | 2.19 | 2 | 0.71 |  |
|  | South West | 43 | 14.0% | 32.6% | 53.5% | 2.4 | 3 | 0.728 |  |
|  | Scotland | 42 | 21.4% | 31.0% | 47.6% | 2.26 | 2 | 0.798 |  |
|  | East of England | 35 | 25.7% | 40.0% | 34.3% | 2.09 | 2 | 0.781 |  |
|  | Yorkshire & the Humber | 46 | 15.2% | 45.7% | 39.1% | 2.24 | 2 | 0.705 |  |
| IMD  Decile (England only) | 1 | 26 | 15.4% | 46.2% | 38.5% | 2.23 | 2 | 0.71 | 0.54 (Medium), 0.169 |
|  | 2 | 40 | 30.0% | 27.5% | 42.5% | 2.13 | 2 | 0.853 |  |
|  | 3 | 45 | 8.9% | 51.1% | 40.0% | 2.31 | 2 | 0.633 |  |
|  | 4 | 50 | 16.0% | 52.0% | 32.0% | 2.16 | 2 | 0.681 |  |
|  | 5 | 58 | 25.9% | 32.8% | 41.4% | 2.16 | 2 | 0.812 |  |
|  | 6 | 44 | 15.9% | 45.5% | 38.6% | 2.23 | 2 | 0.711 |  |
|  | 7 | 49 | 14.3% | 49.0% | 36.7% | 2.22 | 2 | 0.685 |  |
|  | 8 | 51 | 9.8% | 45.1% | 45.1% | 2.35 | 2 | 0.658 |  |
|  | 9 | 40 | 32.5% | 37.5% | 30.0% | 1.98 | 2 | 0.8 |  |
|  | 10 | 47 | 17.0% | 51.1% | 31.9% | 2.15 | 2 | 0.691 |  |

* P < 0.05

**Table S3:** Descriptive statistics (row percentages, means, and medians) for the question “Did you receive information about the approaching death?”. *P*-values are from chi-squared analysis of a contingency table. Effect sizes are estimated from Cohen’s *h*, where *h* = 0.2: small effect, *h* = 0.5: medium effect, *h* ≥ 0.8: large effect.

|  | | *n* | Very well supported | Fairly well supported | A little bit supported | Not at all supported | Mean | Median | SD | *h* (Effect Size),  *P* |
| --- | --- | --- | --- | --- | --- | --- | --- | --- | --- | --- |
| Highest qualification | None / GCSEs | 95 | 9.5% | 9.5% | 27.4% | 53.7% | 3.25 | 4.00 | 0.98 | 0.34 (Small /Medium),  0.028* |
|  | A-level etc. | 122 | 18.0% | 15.6% | 20.5% | 45.9% | 2.94 | 3.00 | 1.16 |  |
|  | HND / Degree etc. | 371 | 17.3% | 20.5% | 23.5% | 38.8% | 2.84 | 3.00 | 1.12 |  |
| Where did they die? | Hospital | 358 | 15.1% | 17.3% | 23.2% | 44.4% | 2.97 | 3.00 | 1.11 | 0.93 (Large),  0.003* |
|  | Home | 113 | 23.0% | 16.8% | 26.5% | 33.6% | 2.71 | 3.00 | 1.16 |  |
|  | Hospice | 33 | 30.3% | 30.3% | 24.2% | 15.2% | 2.24 | 2.00 | 1.06 |  |
|  | Care home | 78 | 5.1% | 16.7% | 20.5% | 57.7% | 3.31 | 4.00 | 0.93 |  |
|  | Other / Don’t Know | 7 | 14.3% | 14.3% | 28.6% | 42.9% | 3.00 | 3.00 | 1.16 |  |
| Cause of death | COVID (Confirmed or Suspected) | 269 | 9.3% | 12.3% | 24.9% | 53.5% | 3.23 | 4.00 | 0.99 | 0.45 (Medium),  <0.001* |
|  | Non-Covid | 321 | 21.8% | 22.4% | 22.4% | 33.3% | 2.67 | 3.00 | 1.15 |  |
| Gender Identity | Male | 61 | 16.4% | 23.0% | 23.0% | 37.7% | 2.82 | 3.00 | 1.12 | 0.06 (Small),  0.892 |
|  | Female | 526 | 16.0% | 17.1% | 23.6% | 43.3% | 2.94 | 3.00 | 1.12 |  |
|  | Other | 3 | 0.0% | 33.3% | 33.3% | 33.3% | 3.00 | 3.00 | 1.00 |  |
| Ethnicity | Non-BAME | 561 | 15.7% | 18.4% | 23.5% | 42.4% | 2.93 | 3.00 | 1.11 | Weak,  0.476 |
|  | BAME | 28 | 21.4% | 7.1% | 25.0% | 46.4% | 2.96 | 3.00 | 1.20 |  |
| Who was it that died? | Partners | 145 | 21.4% | 19.3% | 22.8% | 36.6% | 2.98 | 3.00 | 1.11 | 0.43 (Medium),  0.494 |
|  | Parents | 369 | 15.4% | 15.7% | 23.8% | 45.0% | 3.00 | 3.00 | 0.98 |  |
|  | Grandparents | 24 | 4.2% | 33.3% | 20.8% | 41.7% | 3.27 | 3.00 | 0.80 |  |
|  | Sibling | 15 | 0.0% | 20.0% | 33.3% | 46.7% | 2.92 | 3.00 | 1.17 |  |
|  | Child | 12 | 16.7% | 16.7% | 25.0% | 41.7% | 2.87 | 3.00 | 1.10 |  |
|  | Other family member | 23 | 13.0% | 26.1% | 21.7% | 39.1% | 3.00 | 4.00 | 1.73 |  |
|  | Colleague or friend | 3 | ─ | ─ | ─ | ─ |  |  |  |  |
| Religious beliefs | Yes | 286 | 18.9% | 18.2% | 19.9% | 43.0% | 2.87 | 3.00 | 1.16 | 0.19 (Small),  0.038* |
|  | No | 252 | 12.3% | 16.7% | 29.0% | 42.1% | 3.01 | 3.00 | 1.04 |  |
| Was the death expected? | Yes | 94 | 28.7% | 20.2% | 28.7% | 22.3% | 2.45 | 3.00 | 1.13 | 0.54 (Medium),  <0.001* |
|  | No | 458 | 12.7% | 16.8% | 22.9% | 47.6% | 3.05 | 3.00 | 1.07 |  |
| Region of the UK | Northern Ireland | 19 | 26.3% | 10.5% | 26.3% | 36.8% | 2.74 | 3 | 1.24 | 0.66 (Medium / Large),  0.034* |
|  | Wales | 50 | 8.0% | 14.0% | 28.0% | 50.0% | 3.2 | 3.5 | 0.969 |  |
|  | West Midlands | 48 | 16.7% | 16.7% | 8.3% | 58.3% | 3.08 | 4 | 1.2 |  |
|  | North East | 31 | 9.7% | 35.5% | 25.8% | 29.0% | 2.74 | 3 | 0.999 |  |
|  | Greater London | 51 | 23.5% | 19.6% | 25.5% | 31.4% | 2.65 | 3 | 1.163 |  |
|  | South East | 62 | 14.5% | 12.9% | 21.0% | 51.6% | 3.1 | 4 | 1.112 |  |
|  | North West | 84 | 17.9% | 17.9% | 25.0% | 39.3% | 2.86 | 3 | 1.132 |  |
|  | East Midlands | 34 | 17.6% | 11.8% | 32.4% | 38.2% | 2.91 | 3 | 1.111 |  |
|  | South West | 46 | 19.6% | 17.4% | 21.7% | 41.3% | 2.85 | 3 | 1.173 |  |
|  | Scotland | 45 | 26.7% | 28.9% | 13.3% | 31.1% | 2.49 | 2 | 1.199 |  |
|  | East of England | 35 | 2.9% | 8.6% | 31.4% | 57.1% | 3.43 | 4 | 0.778 |  |
|  | Yorkshire & the Humber | 43 | 14.0% | 14.0% | 32.6% | 39.5% | 2.98 | 3 | 1.058 |  |
| IMD  Decile (England only) | 1 | 24 | 12.5% | 20.8% | 16.7% | 50.0% | 3.04 | 3.5 | 1.122 | 0.47 (Medium),  0.941 |
|  | 2 | 39 | 17.9% | 17.9% | 25.6% | 38.5% | 2.85 | 3 | 1.136 |  |
|  | 3 | 42 | 16.7% | 14.3% | 38.1% | 31.0% | 2.83 | 3 | 1.057 |  |
|  | 4 | 45 | 11.1% | 17.8% | 26.7% | 44.4% | 3.04 | 3 | 1.043 |  |
|  | 5 | 51 | 21.6% | 17.6% | 21.6% | 39.2% | 2.78 | 3 | 1.189 |  |
|  | 6 | 44 | 9.1% | 15.9% | 22.7% | 52.3% | 3.18 | 4 | 1.018 |  |
|  | 7 | 50 | 20.0% | 20.0% | 18.0% | 42.0% | 2.82 | 3 | 1.19 |  |
|  | 8 | 45 | 22.2% | 17.8% | 22.2% | 37.8% | 2.76 | 3 | 1.19 |  |
|  | 9 | 39 | 12.8% | 15.4% | 30.8% | 41.0% | 3 | 3 | 1.051 |  |
|  | 10 | 42 | 11.9% | 16.7% | 19.0% | 52.4% | 3.12 | 4 | 1.087 |  |

* P < 0.05

**Table S4:** Descriptive statistics (row percentages, means, and medians) for the question “Did you feel well supported by the healthcare professionals immediately after the death of your loved one?”. *P*-values are from chi-squared analysis of a contingency table. Effect sizes are estimated from Cohen’s *h*, where *h* = 0.2: small effect, *h* = 0.5: medium effect, *h* ≥ 0.8: large effect.

|  | | *n* | Yes | No | *h* (Effect Size),  *P* |
| --- | --- | --- | --- | --- | --- |
| Highest qualification | None / GCSEs | 93 | 31.2% | 68.8% | 0.43 (Medium),  0.011* |
|  | A-level etc. | 117 | 41.0% | 59.0% |  |
|  | HND / Degree etc. | 360 | 48.1% | 51.9% |  |
| Where did they die? | Hospital | 359 | 42.3% | 57.7% | 0.51 (Medium),  0.216 |
|  | Home | 95 | 44.2% | 55.8% |  |
|  | Hospice | 33 | 63.6% | 36.4% |  |
|  | Care home | 76 | 42.1% | 57.9% |  |
|  | Other / Don’t Know | 8 | 50.0% | 50.0% |  |
| Cause of death | COVID (Confirmed or Suspected) | 269 | 40.9% | 59.1% | 0.13 (Small), 0.206 |
|  | Non-Covid | 303 | 46.2% | 53.8% |  |
| Gender Identity | Male | 60 | 43.3% | 56.7% | 0.01 (None / Small), 0.614 |
|  | Female | 510 | 43.9% | 56.1% |  |
|  | Other | 2 | 0.0% | 100.0% |  |
| Ethnicity | Non-BAME | 543 | 44.4% | 55.6% | 0.31 (Small),  0.243 |
|  | BAME | 28 | 32.1% | 67.9% |  |
| Who was it that died? | Partners | 142 | 52.8% | 47.2% | 0.48 (Medium),  0.353 |
|  | Parents | 355 | 41.1% | 58.9% |  |
|  | Grandparent | 24 | 37.5% | 62.5% |  |
|  | Sibling | 13 | 38.5% | 61.5% |  |
|  | Child | 11 | 45.5% | 54.5% |  |
|  | Other family member | 22 | 40.9% | 59.1% |  |
|  | Colleague or friend | 6 | 33.3% | 66.7% |  |
| Religious beliefs | Yes | 280 | 45.4% | 54.6% | 0.07 (None / Small),  0.537 |
|  | No | 243 | 42.4% | 57.6% |  |
| Was the death expected? | Yes | 88 | 56.8% | 43.2% | 0.36 (Small / Medium), 0.009* |
|  | No | 451 | 41.2% | 58.8% |  |
| Region of the UK | Northern Ireland | 19 | 68.4% | 31.6% | 0.90 (Large),  0.36 |
|  | Wales | 49 | 46.9% | 53.1% |  |
|  | West Midlands | 48 | 37.5% | 62.5% |  |
|  | North East | 30 | 50.0% | 50.0% |  |
|  | Greater London | 52 | 44.2% | 55.8% |  |
|  | South East | 61 | 44.3% | 55.7% |  |
|  | North West | 83 | 47.0% | 53.0% |  |
|  | East Midlands | 33 | 39.4% | 60.6% |  |
|  | South West | 41 | 39.0% | 61.0% |  |
|  | Scotland | 45 | 55.6% | 44.4% |  |
|  | East of England | 33 | 30.3% | 69.7% |  |
|  | Yorkshire & the Humber | 42 | 40.5% | 59.5% |  |
| IMD  Decile (England only) | 1 | 23 | 30.4% | 69.6% | 0.64 (Medium / Large),  0.488 |
|  | 2 | 39 | 46.2% | 53.8% |  |
|  | 3 | 41 | 46.3% | 53.7% |  |
|  | 4 | 44 | 31.8% | 68.2% |  |
|  | 5 | 52 | 44.2% | 55.8% |  |
|  | 6 | 42 | 42.9% | 57.1% |  |
|  | 7 | 48 | 45.8% | 54.2% |  |
|  | 8 | 44 | 34.1% | 65.9% |  |
|  | 9 | 37 | 56.8% | 43.2% |  |
|  | 10 | 41 | 39.0% | 61.0% |  |

* P < 0.05

**Table S5:** Descriptive statistics (row percentages) for the question “Were you contacted again by the hospital or care provider following their death?”. *P*-values are from chi-squared analysis of a contingency table. Effect sizes are estimated from Cohen’s *h*, where *h* = 0.2: small effect, *h* = 0.5: medium effect, *h* ≥ 0.8: large effect.

|  | | *n* | Yes | No | *h* (Effect Size),  *P* |
| --- | --- | --- | --- | --- | --- |
| Highest qualification | None / GCSEs | 108 | 21.3% | 78.7% | 0.08 (Small)  0.848 |
|  | A-level etc. | 132 | 19.7% | 80.3% |  |
|  | HND / Degree etc. | 468 | 22.0% | 78.0% |  |
| Where did they die? | Hospital | 410 | 25.9% | 74.1% | 1.47 (Large),  <0.001* |
|  | Home | 158 | 15.2% | 84.8% |  |
|  | Hospice | 37 | 48.6% | 51.4% |  |
|  | Care home | 91 | 4.4% | 95.6% |  |
|  | Other / don’t know | 13 | 7.7% | 92.3% |  |
| Cause of death | COVID (Confirmed or Suspected) | 311 | 17.7% | 82.3% | 0.22 (Small),  0.028* |
|  | Non-Covid | 399 | 24.6% | 75.4% |  |
| Gender Identity | Male | 74 | 27.0% | 73.0% | 0.19 (Small),  0.529 |
|  | Female | 628 | 21.0% | 79.0% |  |
|  | Other | 7 | 14.3% | 85.7% |  |
| Ethnicity | Non-BAME | 676 | 21.9% | 78.1% | 0.23 (Small),  0.399 |
|  | BAME | 33 | 15.2% | 84.8% |  |
| Who was it that died? | Partners | 152 | 34.9% | 65.1% | 1.17 (Large),  <0.001* |
|  | Parents | 395 | 20.8% | 79.2% |  |
|  | Grandparent | 54 | 9.3% | 90.7% |  |
|  | Sibling | 23 | 8.7% | 91.3% |  |
|  | Child | 15 | 33.3% | 66.7% |  |
|  | Other family member | 46 | 10.9% | 89.1% |  |
|  | Colleague or friend | 26 | 3.8% | 96.2% |  |
| Religious beliefs | Yes | 340 | 21.5% | 78.5% | 0.03 (None / Small),  0.849 |
|  | No | 301 | 22.3% | 77.7% |  |
| Was the death expected? | Yes | 113 | 24.8% | 75.2% | 0.12 (Small),  0.382 |
|  | No | 552 | 21.0% | 79.0% |  |
| Region of the UK | Northern Ireland | 26 | 15.4% | 84.6% | 0.64 (Medium / Large),  0.283 |
|  | Wales | 63 | 14.3% | 85.7% |  |
|  | West Midlands | 52 | 17.3% | 82.7% |  |
|  | North East | 40 | 12.5% | 87.5% |  |
|  | Greater London | 68 | 27.9% | 72.1% |  |
|  | South East | 78 | 21.8% | 78.2% |  |
|  | North West | 95 | 24.2% | 75.8% |  |
|  | East Midlands | 39 | 30.8% | 69.2% |  |
|  | South West | 51 | 17.6% | 82.4% |  |
|  | Scotland | 53 | 32.1% | 67.9% |  |
|  | East of England | 39 | 25.6% | 74.4% |  |
|  | Yorkshire & the Humber | 55 | 20.0% | 80.0% |  |
| IMD  Decile (England only) | 1 | 26 | 34.6% | 65.4% | 0.50 (Medium),  0.805 |
|  | 2 | 45 | 24.4% | 75.6% |  |
|  | 3 | 49 | 20.4% | 79.6% |  |
|  | 4 | 52 | 26.9% | 73.1% |  |
|  | 5 | 64 | 21.9% | 78.1% |  |
|  | 6 | 52 | 28.8% | 71.2% |  |
|  | 7 | 58 | 20.7% | 79.3% |  |
|  | 8 | 57 | 19.3% | 80.7% |  |
|  | 9 | 46 | 19.6% | 80.4% |  |
|  | 10 | 50 | 18.0% | 82.0% |  |

* P < 0.05

**Table S6:** Descriptive statistics for the question “Did they provide information about bereavement support services? – Yes (at the time of death)”. *P*-values are from chi-squared analysis of a contingency table. Effect sizes are estimated from Cohen’s *h*, where *h* = 0.2: small effect, *h* = 0.5: medium effect, *h* ≥ 0.8: large effect.

|  | | *n* | Yes | No | *h* (Effect Size),  *P* |
| --- | --- | --- | --- | --- | --- |
| Highest qualification | None / GCSEs | 108 | 17.6% | 82.4% | 0.27 (Small),  0.09 |
|  | A-level etc. | 132 | 21.2% | 78.8% |  |
|  | HND / Degree etc. | 468 | 13.7% | 86.3% |  |
| Where did they die? | Hospital | 410 | 17.8% | 82.2% | 1.27 (Large),  <0.001* |
|  | Home | 158 | 11.4% | 88.6% |  |
|  | Hospice | 37 | 40.5% | 59.5% |  |
|  | Care home | 91 | 4.4% | 95.6% |  |
|  | Other / DON’T KNOW | 13 | 7.7% | 92.3% |  |
| Cause of death | COVID (Confirmed or Suspected) | 311 | 16.7% | 83.3% | 0.07 (None / Small),  0.532 |
|  | Non-Covid | 399 | 14.8% | 85.2% |  |
| Gender Identity | Male | 74 | 16.2% | 83.8% | 0.02 (None),  0.615 |
|  | Female | 628 | 15.8% | 84.2% |  |
|  | Other | 7 | 0.0% | 100.0% |  |
| Ethnicity | Non-BAME | 676 | 15.8% | 84.2% | 0.15 (Small),  0.638 |
|  | BAME | 33 | 12.1% | 87.9% |  |
| Who was it that died? | Partners | 152 | 23.7% | 76.3% | 1.50 (Large),  0.09 |
|  | Parents | 395 | 15.2% | 84.8% |  |
|  | Grandparents | 54 | 5.6% | 94.4% |  |
|  | Sibling | 23 | 21.7% | 78.3% |  |
|  | Child | 15 | 26.7% | 73.3% |  |
|  | Other family member | 46 | 6.5% | 93.5% |  |
|  | Colleague or friend | 26 | 0.0% | 100.0% |  |
| Religious beliefs | Yes | 340 | 18.2% | 81.8% | 0.18 (Small),  0.104 |
|  | No | 301 | 13.3% | 86.7% |  |
| Was the death expected? | Yes | 113 | 19.5% | 80.5% | 0.15 (Small),  0.323 |
|  | No | 552 | 15.2% | 84.8% |  |
| Region of the UK | Northern Ireland | 26 | 19.2% | 80.8% | 0.59 (Medium),  0.342 |
|  | Wales | 63 | 7.9% | 92.1% |  |
|  | West Midlands | 52 | 19.2% | 80.8% |  |
|  | North East | 40 | 17.5% | 82.5% |  |
|  | Greater London | 68 | 14.7% | 85.3% |  |
|  | South East | 78 | 11.5% | 88.5% |  |
|  | North West | 95 | 23.2% | 76.8% |  |
|  | East Midlands | 39 | 23.1% | 76.9% |  |
|  | South West | 51 | 21.6% | 78.4% |  |
|  | Scotland | 53 | 11.3% | 88.7% |  |
|  | East of England | 39 | 17.9% | 82.1% |  |
|  | Yorkshire & the Humber | 55 | 12.7% | 87.3% |  |
| IMD  Decile (England only) | 1 | 26 | 19.2% | 80.8% | 0.36 (Small / Medium),  0.908 |
|  | 2 | 45 | 15.6% | 84.4% |  |
|  | 3 | 49 | 18.4% | 81.6% |  |
|  | 4 | 52 | 15.4% | 84.6% |  |
|  | 5 | 64 | 20.3% | 79.7% |  |
|  | 6 | 52 | 17.3% | 82.7% |  |
|  | 7 | 58 | 15.5% | 84.5% |  |
|  | 8 | 57 | 24.6% | 75.4% |  |
|  | 9 | 46 | 23.9% | 76.1% |  |
|  | 10 | 50 | 14.0% | 86.0% |  |

* P < 0.05

**Table S7:** Descriptive statistics (row percentages) for the question “Did they provide information about bereavement support services? – Yes (during a follow-up call)”. *P*-values are from chi-squared analysis of a contingency table. Effect sizes are estimated from Cohen’s *h*, where *h* = 0.2: small effect, *h* = 0.5: medium effect, *h* ≥ 0.8: large effect*.*

|  | | Simple Logistic Regression | | | Generalised Linear Model | | |
| --- | --- | --- | --- | --- | --- | --- | --- |
|  |  | OR | 95% LCI | 95% UCI | OR | 95% LCI | 95% UCI |
| Who was it that died? | Partners | Reference Class | | | | | |
|  | Parents | 1.571 | 1.076 | 2.294 | 1.310 | 0.787 | 2.180 |
|  | Grandparent | 4.944 | 2.411 | 10.139 | 9.332 | 2.033 | 42.841 |
|  | Sibling | 2.649 | 1.059 | 6.625 | 6.387 | 1.150 | 35.473 |
|  | Child | 0.706 | 0.230 | 2.167 | 1.357 | 0.217 | 8.483 |
|  | Other family member | 5.808 | 2.618 | 12.883 | 7.716 | 1.860 | 32.014 |
|  | Colleague or friend | 2.260 | 0.963 | 5.307 | ─ | ─ | ─ |
| Where did they die? | In hospital | Reference Class | | | | | |
|  | In their home | 0.242 | 0.163 | 0.359 | 0.190 | 0.087 | 0.417 |
|  | In a hospice | 0.435 | 0.220 | 0.859 | 0.887 | 0.341 | 2.310 |
|  | In a care home | 1.053 | 0.655 | 1.693 | 1.181 | 0.622 | 2.242 |
|  | Other / Don’t Know | 0.104 | 0.023 | 0.475 | 0.864 | 0.080 | 9.339 |
| Cause of Death | COVID (Confirmed or Suspected) | Reference Class | | | | | |
|  | Non-Covid | 0.318 | 0.233 | 0.435 | 0.601 | 0.361 | 1.000 |
| Highest qualification | None / GCSEs | Reference Class | | | | | |
|  | A-level / apprenticeship / ONC | 0.782 | 0.468 | 1.306 | 1.217 | 0.609 | 2.431 |
|  | HND / University Degree / Postgraduate (etc) | 0.833 | 0.546 | 1.273 | 1.111 | 0.610 | 2.022 |
| Gender | Male | Reference Class | | | | | |
|  | Female | 1.188 | 0.734 | 1.924 | 0.640 | 0.311 | 1.319 |
|  | Other | 6.000 | 0.688 | 52.313 | 0.000 | 0.000 | ─ |
| Did you expect your loved one to die around this time? | Yes | Reference Class | | | | | |
|  | No | 3.288 | 2.131 | 5.072 | 1.404 | 0.661 | 2.982 |
|  | Don’t Know | 2.127 | 1.037 | 4.365 | 0.880 | 0.292 | 2.652 |

Note: The symbol “─” is given when results of simple linear regression or of the GLM became unreliable due to small sample sizes.

**Table S8:** Results for the Odds Ratio (OR) and associated 95% confidence intervals from simple logistic regression and a mixed-effects generalised linear model for the item “Unable to visit them prior to their death.”

|  | | Simple Logistic Regression | | | Generalised Linear Model | | |
| --- | --- | --- | --- | --- | --- | --- | --- |
|  |  | OR | 95% LCI | 95% UCI | OR | 95% LCI | 95% UCI |
| Who was it that died? | Partners | Reference Class | | | | | |
|  | Parents | 2.020 | 1.383 | 2.950 | 1.587 | 0.958 | 2.627 |
|  | Grandparent | 1.991 | 1.052 | 3.768 | 0.960 | 0.313 | 2.947 |
|  | Sibling | 1.523 | 0.629 | 3.686 | 1.240 | 0.277 | 5.545 |
|  | Child | 0.426 | 0.130 | 1.397 | 0.094 | 0.009 | 0.982 |
|  | Other family member | 1.998 | 1.014 | 3.938 | 1.335 | 0.429 | 4.157 |
|  | Colleague or friend | 0.859 | 0.371 | 1.992 | ─ | ─ | ─ |
| Where did they die? | In hospital | Reference Class | | | | | |
|  | In their home | 0.244 | 0.165 | 0.362 | 0.110 | 0.050 | 0.240 |
|  | In a hospice | 0.391 | 0.198 | 0.773 | 0.557 | 0.226 | 1.371 |
|  | In a care home | 1.710 | 1.008 | 2.901 | 1.390 | 0.700 | 2.757 |
|  | Other / Don’t Know | 0.093 | 0.020 | 0.427 | 0.235 | 0.021 | 2.644 |
| Cause of Death | COVID (Confirmed or Suspected) | Reference Class | | | | | |
|  | Non-Covid | 0.380 | 0.278 | 0.520 | 0.742 | 0.446 | 1.236 |
| Highest qualification | None / GCSEs | Reference Class | | | | | |
|  | A-level / apprenticeship / ONC | 1.802 | 1.071 | 3.031 | 2.431 | 1.198 | 4.935 |
|  | HND / University Degree / Postgraduate (etc) | 1.291 | 0.849 | 1.964 | 1.950 | 1.081 | 3.515 |
| Gender | Male | Reference Class | | | | | |
|  | Female | 1.588 | 0.979 | 2.574 | 1.425 | 0.723 | 2.810 |
|  | Other | 2.786 | 0.508 | 15.281 | ─ | ─ | ─ |
| Did you expect your loved one to die around this time? | Yes | Reference Class | | | | | |
|  | No | 2.995 | 1.964 | 4.568 | 1.434 | 0.699 | 2.941 |
|  | Don’t Know | 2.791 | 1.355 | 5.750 | 1.678 | 0.579 | 4.860 |

Note: The symbol “─” is given when results of simple linear regression or of the GLM became unreliable due to small sample sizes

**Table S9:** Results for the Odds Ratio (OR) and associated 95% confidence intervals from simple logistic regression and a mixed-effects generalised linear model for the item “Limited contact with them in last days of their life.”

|  | | Simple Logistic Regression | | | Generalised Linear Model | | |
| --- | --- | --- | --- | --- | --- | --- | --- |
|  |  | OR | 95% LCI | 95% UCI | OR | 95% LCI | 95% UCI |
| Who was it that died? | Partners | Reference Class | | | | | |
|  | Parents | 1.933 | 1.323 | 2.826 | 1.561 | 0.897 | 2.714 |
|  | Grandparent | 3.593 | 1.756 | 7.353 | 2.142 | 0.577 | 7.948 |
|  | Sibling | 2.909 | 1.088 | 7.778 | 1.843 | 0.397 | 8.561 |
|  | Child | 1.173 | 0.405 | 3.397 | 2.267 | 0.351 | 14.624 |
|  | Other family member | 4.221 | 1.906 | 9.345 | 4.118 | 1.062 | 15.974 |
|  | Colleague or friend | 1.939 | 0.814 | 4.621 | ─ | ─ | ─ |
| Where did they die? | In hospital | Reference Class | | | | | |
|  | In their home | 0.231 | 0.157 | 0.340 | 0.212 | 0.099 | 0.454 |
|  | In a hospice | 0.172 | 0.083 | 0.354 | 0.618 | 0.23 | 1.662 |
|  | In a care home | 1.450 | 0.828 | 2.541 | 2.089 | 0.951 | 4.588 |
|  | Other / Don’t Know | 0.159 | 0.048 | 0.527 | 0.054 | 0 | 16.76 |
| Cause of Death | COVID (Confirmed or Suspected) | Reference Class | | | | | |
|  | Non-Covid | 0.186 | 0.130 | 0.266 | 0.348 | 0.2 | 0.605 |
| Highest qualification | None / GCSEs | Reference Class | | | | | |
|  | A-level / apprenticeship / ONC | 0.787 | 0.456 | 1.360 | 1.181 | 0.526 | 2.654 |
|  | HND / University Degree / Postgraduate (etc) | 0.686 | 0.436 | 1.079 | 1.034 | 0.515 | 2.075 |
| Gender | Male | Reference Class | | | | | |
|  | Female | 1.769 | 1.090 | 2.872 | 1.566 | 0.747 | 3.281 |
|  | Other | 2.368 | 0.432 | 12.991 | ─ | ─ | ─ |
| Did you expect your loved one to die around this time? | Yes | Reference Class | | | | | |
|  | No | 3.818 | 2.506 | 4.568 | 1.101 | 0.523 | 2.316 |
|  | Don’t Know | 1.705 | 0.840 | 5.750 | 0.525 | 0.168 | 1.643 |

Note: The symbol “─” is given when results of simple linear regression or of the GLM became unreliable due to small sample sizes.

**Table S10:** Results for the Odds Ratio (OR) and associated 95% confidence intervals from simple logistic regression and a mixed-effects generalised linear model for the item “Unable to say goodbye as I would have liked.”

|  | | Simple Logistic Regression | | | Generalised Linear Model | | |
| --- | --- | --- | --- | --- | --- | --- | --- |
|  |  | OR | 95% LCI | 95% UCI | OR | 95% LCI | 95% UCI |
| Who was it that died? | Partners | Reference Class | | | | | |
|  | Parents | 0.942 | 0.410 | 2.164 | 1.091 | 0.457 | 2.603 |
|  | Grandparents | 0.544 | 0.170 | 1.743 | 0.545 | 0.104 | 2.853 |
|  | Sibling | 0.370 | 0.091 | 1.512 | 0.596 | 0.065 | 5.484 |
|  | Child | 0.778 | 0.091 | 6.677 | 0.356 | 0.034 | 3.776 |
|  | Other family member | 0.796 | 0.202 | 3.133 | 0.993 | 0.128 | 7.72 |
|  | Colleague or friend | 0.233 | 0.070 | 0.781 | ─ | ─ | ─ |
| Where did they die? | In hospital | Reference Class | | | | | |
|  | In their home | 0.790 | 0.387 | 1.614 | 0.561 | 0.189 | 1.665 |
|  | In a hospice | ─ | ─ | ─ | 2.642 | 0.289 | 24.113 |
|  | In a care home | 0.592 | 0.266 | 1.314 | 0.315 | 0.118 | 0.838 |
|  | Other / Don’t Know | 0.779 | 0.097 | 6.235 | 1.771 | 0.005 | 649.828 |
| Cause of Death | COVID (Confirmed or Suspected) | Reference Class | | | | | |
|  | Non-Covid | 0.523 | 0.275 | 0.995 | 0.66 | 0.273 | 1.592 |
| Highest qualification | None / GCSEs | Reference Class | | | | | |
|  | A-level / apprenticeship / ONC | 2.444 | 0.939 | 6.362 | 1.631 | 0.564 | 4.721 |
|  | HND / University Degree / Postgraduate (etc) | 2.326 | 1.153 | 4.693 | 1.918 | 0.794 | 4.635 |
| Gender | Male | Reference Class | | | | | |
|  | Female | 2.496 | 1.185 | 5.255 | 2.292 | 0.906 | 5.799 |
|  | Other | ─ | ─ | ─ | ─ | ─ | ─ |
| Did you expect your loved one to die around this time? | Yes | Reference Class | | | | | |
|  | No | 2.112 | 1.071 | 4.167 | 1.179 | 0.397 | 3.502 |
|  | Don’t Know | 2.665 | 0.576 | 12.337 | 1.812 | 0.287 | 11.428 |

Note: The symbol “─” is given when results of simple linear regression or of the GLM became unreliable due to small sample sizes

**Table S11:** Results for the Odds Ratio (OR) and associated 95% confidence intervals (i.e., 95% LCI and 95% UCI) from simple logistic regression and a mixed-effects generalised linear model for the item “Restricted funeral arrangements.”

|  | | Simple Logistic Regression | | | Generalised Linear Model | | |
| --- | --- | --- | --- | --- | --- | --- | --- |
|  |  | OR | 95% LCI | 95% UCI | OR | 95% LCI | 95% UCI |
| Who was it that died? | Partners | Reference Class | | | | | |
|  | Parents | 0.421 | 0.266 | 0.666 | 0.292 | 0.158 | 0.539 |
|  | Grandparents | 0.304 | 0.155 | 0.599 | 0.159 | 0.051 | 0.493 |
|  | Sibling | 0.294 | 0.117 | 0.737 | 0.114 | 0.025 | 0.514 |
|  | Child | 0.452 | 0.143 | 1.425 | 0.718 | 0.103 | 5.021 |
|  | Other family member | 0.246 | 0.121 | 0.501 | 0.092 | 0.028 | 0.297 |
|  | Colleague or friend | 0.308 | 0.128 | 0.742 | ─ | ─ | ─ |
| Where did they die? | In hospital | Reference Class | | | | | |
|  | In their home | 0.804 | 0.547 | 1.183 | 1.371 | 0.664 | 2.831 |
|  | In a hospice | 1.412 | 0.648 | 3.079 | 2.734 | 0.952 | 7.852 |
|  | In a care home | 0.798 | 0.496 | 1.284 | 0.81 | 0.426 | 1.541 |
|  | Other / Don’t Know | 0.284 | 0.091 | 0.884 | ─ | ─ | ─ |
| Cause of Death | COVID (Confirmed or Suspected) | Reference Class | | | | | |
|  | Non-Covid | 0.483 | 0.348 | 0.670 | 0.439 | 0.261 | 0.739 |
| Highest qualification | None / GCSEs | Reference Class | | | | | |
|  | A-level / apprenticeship / ONC | 0.959 | 0.558 | 1.648 | 1.052 | 0.523 | 2.115 |
|  | HND / University Degree / Postgraduate (etc) | 0.950 | 0.608 | 1.484 | 1.597 | 0.867 | 2.94 |
| Gender | Male | Reference Class | | | | | |
|  | Female | 1.238 | 0.752 | 2.038 | 1.439 | 0.729 | 2.841 |
|  | Other | 1.522 | 0.276 | 8.378 | ─ | ─ | ─ |
| Did you expect your loved one to die around this time? | Yes | Reference Class | | | | | |
|  | No | 1.543 | 1.017 | 2.340 | 1.211 | 0.598 | 2.454 |
|  | Don’t Know | 0.867 | 0.427 | 1.763 | 0.645 | 0.241 | 1.725 |

Note: The symbol “─” is given when results of simple linear regression or of the GLM became unreliable due to small sample sizes.

**Table S12:** Results for the Odds Ratio (OR) and associated 95% confidence intervals (i.e., 95% LCI and 95% UCI) from simple logistic regression and a mixed-effects generalised linear model for the item “Social isolation and loneliness.”

|  | | Simple Logistic Regression | | | Generalised Linear Model | | |
| --- | --- | --- | --- | --- | --- | --- | --- |
|  |  | OR | 95% LCI | 95% UCI | OR | 95% LCI | 95% UCI |
| Who was it that died? | Partners | Reference Class | | | | | |
|  | Parents | 1.134 | 0.700 | 1.835 | 0.91 | 0.504 | 1.643 |
|  | Grandparents | 0.744 | 0.354 | 1.564 | 0.329 | 0.104 | 1.04 |
|  | Sibling | 0.668 | 0.242 | 1.843 | 0.265 | 0.055 | 1.289 |
|  | Child | 0.648 | 0.193 | 2.183 | 2.068 | 0.207 | 20.613 |
|  | Other family member | 1.120 | 0.472 | 2.655 | 0.948 | 0.234 | 3.842 |
|  | Colleague or friend | 0.445 | 0.180 | 1.099 | ─ | ─ | ─ |
| Where did they die? | In hospital | Reference Class | | | | | |
|  | In their home | 0.749 | 0.476 | 1.178 | 1.111 | 0.51 | 2.422 |
|  | In a hospice | 1.757 | 0.604 | 5.115 | 2.409 | 0.701 | 8.278 |
|  | In a care home | 0.756 | 0.433 | 1.321 | 0.968 | 0.45 | 2.084 |
|  | Other / Don’t Know | 0.341 | 0.108 | 1.072 | 0.206 | 0.025 | 1.671 |
| Cause of Death | COVID (Confirmed or Suspected) | Reference Class | | | | | |
|  | Non-Covid | 0.479 | 0.321 | 0.716 | 0.465 | 0.254 | 0.852 |
| Highest qualification | None / GCSEs | Reference Class | | | | | |
|  | A-level / apprenticeship / ONC | 1.646 | 0.898 | 3.017 | 1.73 | 0.836 | 3.581 |
|  | HND / University Degree / Postgraduate (etc) | 1.811 | 1.116 | 2.937 | 2.833 | 1.504 | 5.337 |
| Gender | Male | Reference Class | | | | | |
|  | Female | 2.101 | 1.232 | 3.580 | 2.116 | 1.059 | 4.227 |
|  | Other | 0.601 | 0.124 | 2.907 | ─ | ─ | ─ |
| Did you expect your loved one to die around this time? | Yes | Reference Class | | | | | |
|  | No | 1.334 | 0.819 | 2.175 | 1.29 | 0.598 | 2.785 |
|  | Don’t Know | 1.129 | 0.480 | 2.656 | 1.092 | 0.351 | 3.396 |

Note: The symbol “─” is given when results of simple linear regression or of the GLM became unreliable due to small sample sizes

**Table S13:** Results for the Odds Ratio (OR) and associated 95% confidence intervals (i.e., 95% LCI and 95% UCI) from simple logistic regression and a mixed-effects generalised linear model for the item “Limited contact with other close relatives or friends.”
